# BMI Deviations from Genetic Prediction Are Associated with Distinct Molecular Signatures in Finnish Population Cohorts

**DOI:** 10.64898/2026.10.21.26366088

**Authors:** Tianyu Zhu, Gabin Drouard, Aino Heikkinen, Mikaela Hukkanen, Teemu Palviainen, Bram J. Berntzen, Leo-Pekka Lyytikäinen, Emma Raitoharju, Kirsi H. Pietiläinen, Terho Lehtimäki, Olli Raitakari, Mika Ala-Korpela, Jaakko Kaprio, Robin Cristofari, Miina Ollikainen

## Abstract

**Objective:** Polygenic risk scores (PRS) estimate genetically predicted BMI, but lifestyle and environmental factors may cause deviations between observed and predicted BMI. We investigated whether such deviations are associated with distinct molecular profiles and whether these differ by direction of deviation.

**Methods:** We analyzed 2,508 adults from the Finnish Twin Cohort and 1,715 from the Young Finns Study with clinical, metabolomic, DNA methylation, and proteomic data. PRS-informed residual BMI (rBMI) quantified deviations between observed and genetically predicted BMI. Associations between rBMI and multi-omic profiles were assessed using linear mixed-effects models with a hinge term to compare effects above *vs.* below genetic prediction. Twin analyses further accounted for shared genetic and environmental factors.

**Results:** rBMI is associated with distinct cardiometabolic and molecular profiles depending on whether BMI is above or below genetic prediction. When BMI exceeds genetic prediction, rBMI shows stronger associations with inflammatory and cardiovascular markers. In contrast, several metabolites and plasma proteins show stronger associations when BMI is below genetic prediction. Epigenetic aging increases with higher rBMI regardless of direction. Similar patterns are observed in BMI-discordant twin pairs.

**Conclusion:** Deviations from genetically predicted BMI are associated with distinct clinical and molecular signatures beyond absolute BMI alone.

## Introduction

Body mass index (BMI), calculated as weight in kilograms divided by height in meters squared, is a widely used measure of body size and is influenced by both environmental and genetic factors. Twin, family and adoption studies estimate BMI heritability at 40%–70% (1,2), although this decreases with age as environmental exposures accumulate (3,4). Genome-wide association studies (GWAS) have identified numerous BMI-associated loci (5), and recent multi-ancestry polygenic risk scores (PRS) explain up to 17.6% of BMI variation in European-ancestry populations (6), providing an estimate of genetic susceptibility to obesity.

Interestingly, individuals with similar BMI often differ markedly in metabolic health, which may partly depend on whether BMI aligns with an individual’s genetic predisposition. For example, individuals with obesity and a higher BMI PRS have been reported to have a lower risk of cardiovascular disease (CVD) than those with a lower PRS (7). Likewise, BMI that deviates from genetically predicted values, either above or below expectation, has been associated with increased mortality (8). These findings suggest that individuals may have genetically determined ranges of tolerated adiposity, while deviations from this range may be harmful. However, the molecular correlates of such deviations remains largely unknown.

Multi-omics studies have linked BMI to widespread changes in DNA methylation (9,10), circulating metabolites (11), plasma proteins (12), and biological aging measured by epigenetic and proteomic clocks (13–18). However, conventional population studies cannot fully distinguish genetic from environmental contributions to these molecular signatures. Twin studies, particularly BMI-discordant monozygotic (MZ) pairs, provide a powerful complementary approach by controlling for shared genetics and early-life environment, enabling identification of molecular changes associated with BMI independent of these factors (19).

In this study, we leveraged multi-omics data from the Finnish Twin Cohort (FTC) (20–22) and Young Finns Study (YFS) (23–25) to investigate the molecular correlates of deviation from genetically predicted BMI. Building on previous work (26), we derived a PRS-informed residual BMI (rBMI) to quantify the difference between observed and genetically predicted BMI. We first applied population-level hinge regression to determine whether associations between rBMI and clinical, metabolomic, epigenetic, and proteomic traits differ when BMI is above *vs.* below genetic prediction. We then evaluated these associations within BMI-discordant twin pairs, stratified according to whether the higher-BMI (Group A) or lower-BMI (Group B) co-twin deviated from their genetic prediction. We aimed to determine whether deviations from genetically predicted BMI are associated with distinct molecular signatures and whether these differ depending on the direction of deviation.

## Methods

### Study cohorts

The Finnish Twin Cohort (FTC) is a population-based longitudinal study investigating genetic and environmental influences on health and behavior, comprising the Older Twin Cohort (21), FinnTwin12 (FT12) (22) and FinnTwin16 (FT16) (20). BMI was obtained either from self-reported height and weight at the time of blood sampling, previously shown to be reliable (27), or from clinically measured values in a metabolic sub-cohort (28). DNA methylation, metabolomic, and proteomic data were available for subsets of participants (Supplementary Methods).

The Cardiovascular Risk in Young Finns Study (YFS) is a prospective Finnish cohort initiated in 1980 that includes repeated follow-up examinations from childhood into adulthood (23,25). Serum metabolomics were measured using a high-throughput NMR platform (29,30), and DNA methylation data were available for a subset of participants (Supplementary Methods).

Participants with BMI, age, sex, and PRS data and at least one omics dataset were included. Analyses were restricted to adults (> 20 years) without diabetes and with BMI ranging 16 and 45 kg/m^2^, yielding 2,508 FTC participants and 1,715 YFS participants.

### Polygenic risk score and residual BMI

The BMI PRS was derived from 996,919 common SNPs using previously described methods (31), based on HapMap3 SNPs and independent Finnish linkage disequilibrium reference data (32). The same procedure was applied to FTC and YFS.

Residual BMI (rBMI) was calculated separately in FTC and YFS, as the residual from a linear regression of BMI on BMI PRS, adjusted for age and sex. Positive rBMI indicates BMI above genetic prediction, whereas negative rBMI indicates BMI below genetic prediction.

### Population-level analyses

To determine whether rBMI has differential effects on the outcome (clinical measurements, metabolomics, proteomics, DNAm, or epigenetic clocks) when it is above versus below zero (i.e., when the observed BMI is higher or lower than the genetically predicted BMI), we implemented a hinge regression model on FTC and YFS, with family ID as random intercept to adjust for twin relatedness in FTC. This approach estimates separate slopes on either side of the knot at rBMI = 0. The model was specified as:

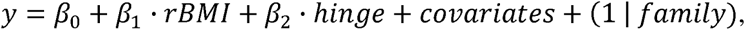

where the hinge term is defined as:

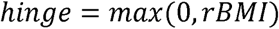

In this parameterization,*β_1_* represents the slope for rBMI values below zero (below-prediction), whereas *β_1_*+*β_2_* corresponds to the slope for rBMI values above zero (above-prediction). The statistical significance of *β_2_* indicates whether the two slopes differ significantly from each other. P values were adjusted for multiple testing using the Benjamini-Hochberg false discovery rate (FDR) procedure, applied separately within each omics layer in each cohort; unless otherwise specified, nominal P values are reported in the main results. Details of hinge regression analyses for each omics layer are described in the Supplementary Methods.

### BMI-discordant twin analyses

The population standard deviation (SD) of rBMI in FTC was 4.2 kg/m^2^. Individuals with |rBMI| ≥ 2.1 kg/m^2^ (0.5 SD) were classified as deviating from genetic prediction. Twin pairs with an intrapair BMI difference (ΔBMI) > 3 kg/m² were classified as BMI-discordant. Based on these criteria, two groups were defined as **(A) *<u>A</u>****bove-prediction* (one twin follows prediction, |rBMI| ≤ 2.1 kg/m^2^, one twin above prediction, rBMI > 2.1 kg/m^2^ and ΔBMI > 3), N=122 pairs (58 monozygotic, 64 dizygotic) and **(B) *<u>B</u>****elow-prediction* (one twin follows prediction, |rBMI| ≤ 2.1 kg/m^2^, one twin below prediction, rBMI < -2.1 kg/m^2^ and ΔBMI > 3), N=109 pairs (39 monozygotic, 70 dizygotic) - see Fig. 1a. We evaluated a set of different thresholds to group the twins, as ±0.1 SD, ±0.25 SD, ±0.5 SD, ±0.75 SD, ±1 SD, ±1.25 SD, ±1.5 SD of rBMI. We selected ±0.5 SD for primary analyses as it resulted in balanced and adequate samples in the two groups (Fig. S1).

**Figure 1.**
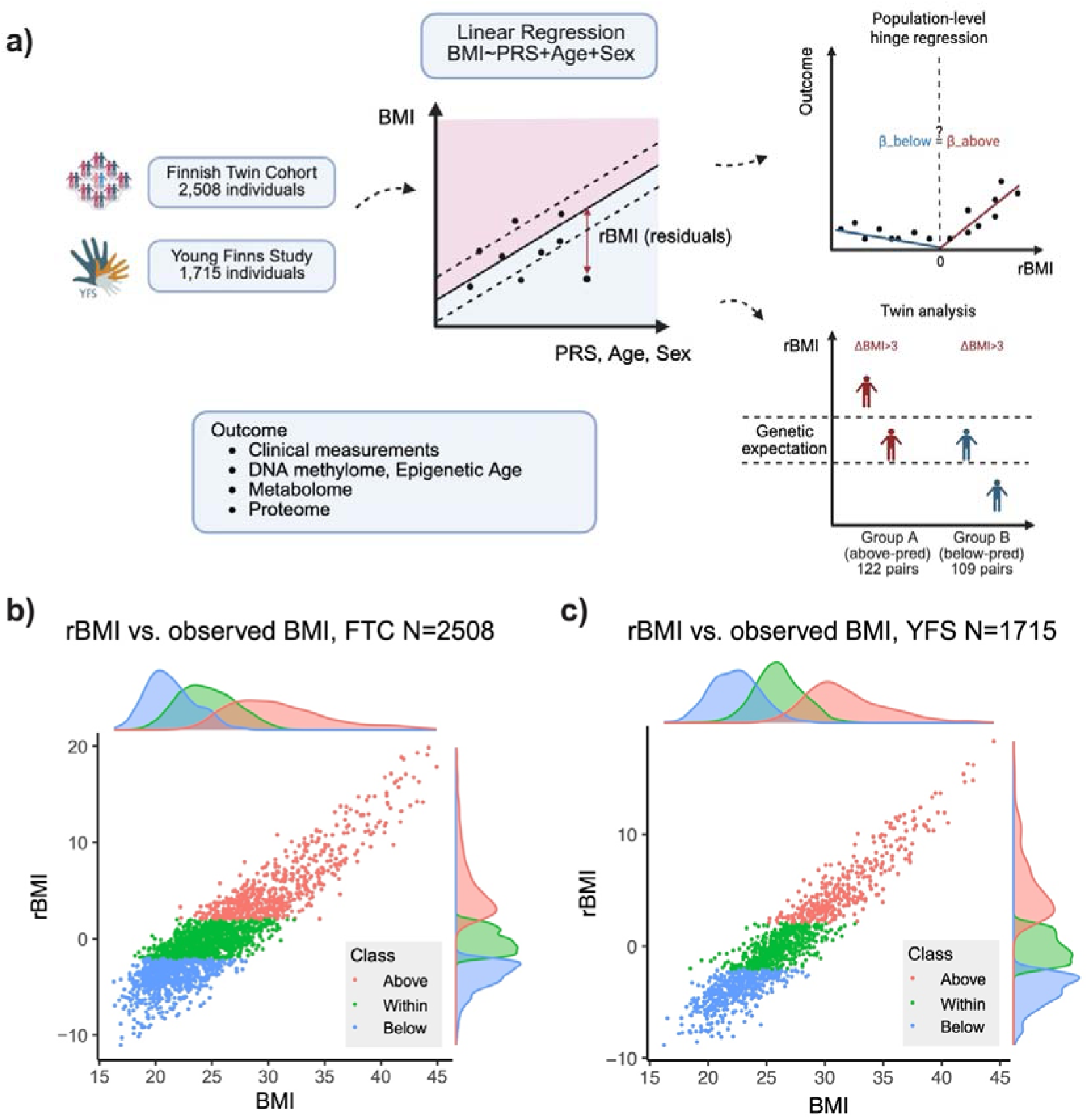
Study overview. **a)** Study Workflow. PRS-informed residual BMI (rBMI) was derived as the residual from linear regression of observed BMI on the polygenic risk score (PRS) for BMI, adjusted for age and sex. We first examined population-level associations between rBMI and multi-omic profiles using linear mixed-effect models with a hinge term to assess differential effects above versus below genetically predicted BMI. Then we evaluated them within BMI-discordant twin pairs, stratified based on whether the higher-BMI co-twin exceeded genetic prediction (Group A) or the lower-BMI co-twin fell below genetic prediction (Group B). Clinical, DNA methylation, metabolomic, and proteomic measurements were analyzed as outcomes. Created in BioRender. Zhu, T. (2026) https://BioRender.com/r3evqlk **b)** Scatterplot showing the correlation between rBMI (y-axis) and actual BMI (x-axis) for each sample in FTC. Samples are colored based on whether their rBMI is above (red), within (green), or below (blue) ±0.5 standard deviation from mean (zero). The density of rBMI and actual BMI for each category is shown on the side. **c)** Same as b), but for YFS participants.

We estimated within-pair associations using a linear fixed-effects model (33), treating twin pairs as the fixed-effect dimension, as in (34). For each individual *j* of twin pair *i*, outcome *Y_ij_* (e.g., clinical measurements, metabolite levels, CpG methylation beta values, epigenetic age, or protein levels) was modeled as:

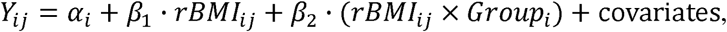

where *α*_i_ denotes the twin-pair-specific intercept that models all factors constant within a pair, including shared genetic and early-life environment. Group indicates the above (Group A) and below (Group B) prediction group described above. Because the Group variable is constant within each-pair, its main effect is absorbed by the fixed effect and not separately estimated. The interaction term *β*_2_ tests whether the within-pair association between rBMI and the outcome differs between Group A and Group B, with Group B as the reference level. Covariates matched those used in the hinge regression described above, excluding age and sex, which are inherently controlled in the same-sex twin design. Analysis was performed using the *feols* function from *fixest* R package (Bergé, 2018), with standard errors clustered by twin family ID to account for residual within-pair correlation.

## Results

### Study design

We analyzed multi-omics data from FTC and YFS to investigate molecular correlates of deviation from genetically predicted BMI (Fig. 1a, Table 1). Residual BMI (rBMI) quantified deviation between observed BMI and BMI predicted by PRS, age and sex (Methods). The PRS explained around 5% and 10% of BMI variance in FTC and YFS, respectively, increasing to 19% and 13% after adjustment for age and sex. Indeed, we observed interactions between BMI and PRS, age, and sex with longitudinal data of 11,428 FTC individuals (Fig. S2).

**Table 1.** Descriptive statistics.

|  | FTC | YFS | twin Group A | twin Group B |
| --- | --- | --- | --- | --- |
| Sample size | 2,508 | 1,715 | 122 pairs | 109 pairs |
| Female sex, N (%) | 1653 (65.9%) | 940 (54.8%) | 85 pairs (69.7%) | 80 pairs (73.4%) |
| Age, mean (SD) | 40.0 (19.6) | 41.9 (5.0) | 45.4 (19.0) | 44.5 (20.1) |
| Higher BMI, mean (SD) |  |  | 31.3 (4.0) | 25.9 (2.3) |
| Lower BMI, mean (SD) |  |  | 25.1 (2.5) | 21.2 (2.0) |
| Higher-Lower BMI, mean (SD) |  |  | 6.2 (3.4) | 4.6 (1.7) |
| Higher-Lower rBMI, mean (SD) |  |  | 5.9 (3.1) | 4.8 (1.4) |

twin Group A: BMI-discordant ( $\Delta\text{BMI}>3$ ) same-sex twin pairs with the higher-BMI co-twin's BMI above genetic prediction and the lower-BMI co-twin's BMI within genetic prediction
twin Group B: BMI-discordant ( $\Delta\text{BMI}>3$ ) same-sex twin pairs with the higher-BMI co-twin's BMI within genetic prediction and the lower-BMI co-twin's BMI below genetic prediction

We combined population-level hinge regression with within-pair analyses of BMI-discordant twins stratified according to whether the higher- or lower-BMI co-twin deviated from genetic prediction (Fig. 1a, Methods). While individuals with a high, intermediate, and low BMI tend to fall within above-, within-, and below-prediction categories respectively, some overlap across different categories was observed (Fig. 1b-c). The rBMI distribution in higher- and lower-BMI co-twin in the two groups are shown in Fig. S3a. The within-pair rBMI difference was slightly larger in Group A than Group B (Wilcoxon rank-sum Test, P=0.036, Table 1, Fig. S3b). The two groups did not differ significantly in age or sex (Table 1, Fig. S3c-d). There was no difference in smoking (proxied by AHRR methylation levels) between higher- and lower-BMI co-twins in either group. However, overall smoking exposure was marginally higher in Group B compared to Group A (Wilcoxon rank-sum test, P = 0.047; Fig. S3e).

### PRS for BMI is associated with clinical and metabolic measurements

We first examined whether the PRS for BMI was associated with obesity-related clinical and metabolic measurements in the FTC (Supplementary Methods). Higher PRS values were significantly associated with elevated C-reactive protein, blood pressure, intra-abdominal fat, waist circumference, fat mass, fat percentage, and gamma-glutamyl transferase (GGT) (Fig. 2a). In metabolomic analyses, higher PRS was additionally associated with increased valine levels, a higher APOB/APOA1 ratio, greater proportions of monounsaturated fatty acids (MUFA), and higher total triglycerides, along with lower HDL cholesterol (Fig. 2b). These associations confirm that PRS for BMI captures biologically relevant variation in metabolic and clinical outcomes in this cohort, in line with known associations with BMI.

**Figure 2.**
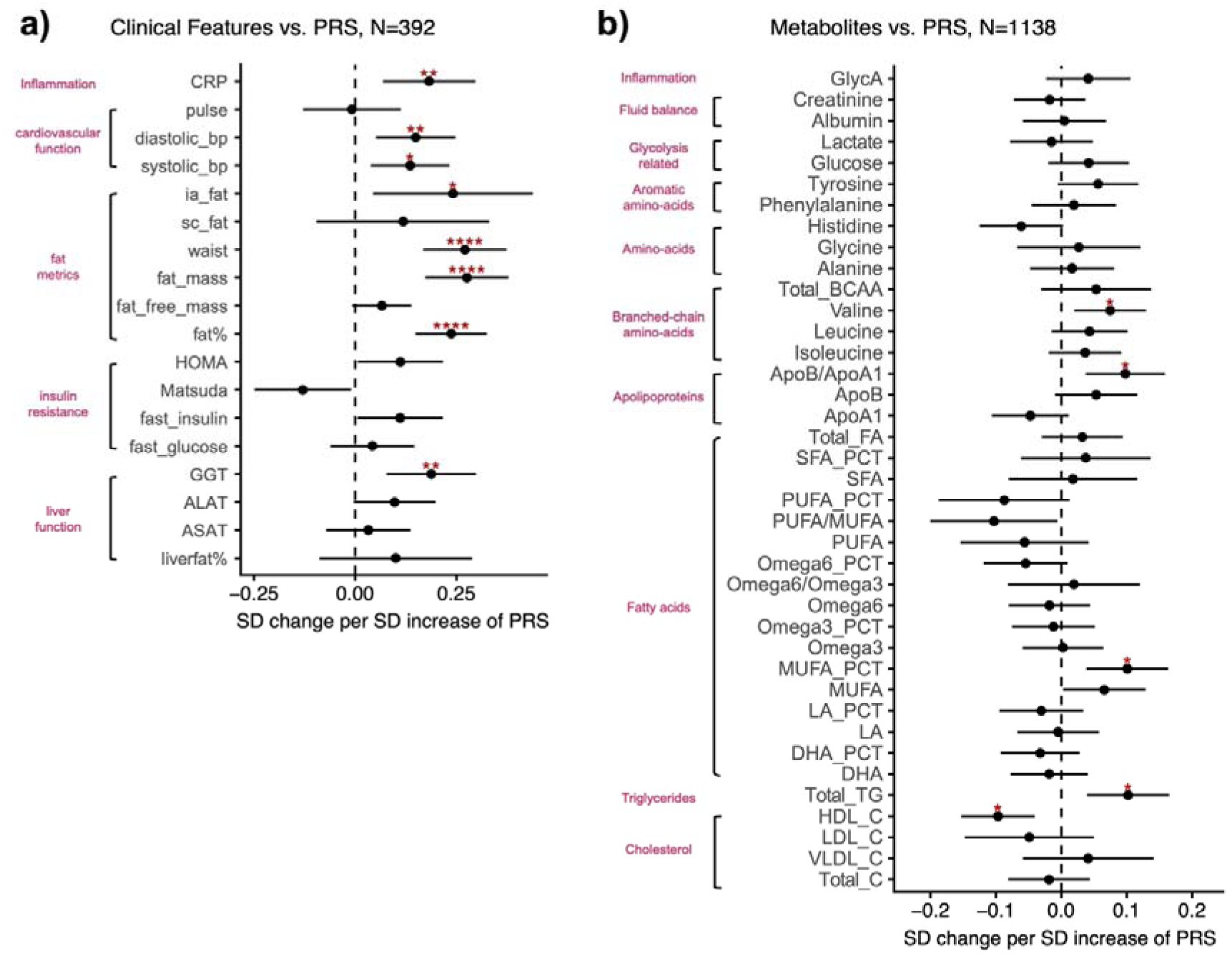
Associations between PRS for BMI and clinical and metabolic measurements in FTC participants. **a)** Forest plots showing the coefficients (dots) with 95% confidence intervals (error bars) derived from linear mixed-effect models fitted in FTC participants. Clinical measurements (y-axis) were regressed on the PRS for BMI (x-axis), adjusting for age and sex as fixed effects and family ID as a random effect. **b)** Same as a), but for 39 clinically validated metabolic biomarkers as described by Julkunen et al. (2023). P-values were FDR-adjusted using the Benjamini-Hochberg false discovery rate (FDR). Significance level: *, FDR<0.05; **, FDR<0.01; ***, FDR<0.001; ****, FDR<0.0001. Abbreviations: CRP, C-reactive protein; bp, blood pressure; ia, intra-abdominal; sc, subcutaneous; HOMA, the homeostatic model assessment used to quantify insulin resistance and beta-cell function; Matsuda, Matsuda index from oral glucose tolerance test; GGT, gamma-glutamyl transferase; ALAT, alanine transaminase; ASAT, aspartate aminotransferase; BCAA, branched-chain amino acid; SFA, saturated fatty acid; PUFA, polyunsaturated fatty acids; MUFA, monounsaturated fatty acids; LA, linoleic acid. _C, cholesterol.

### Differential associations above and below genetically predicted BMI

To identify molecular features showing differential associations depending on the direction of deviation from genetically predicted BMI, we examined differences in rBMI effects above versus below zero using hinge regression models across multiple data layers (Methods and Supplementary Methods).

#### Clinical measurements

In a subset of 392 individuals from the FTC metabolic sub-cohort (28) with obesity-related clinical measurements, several measurements showed differential associations with rBMI depending on whether BMI was above or below genetic prediction (Fig. 3a-b). C-reactive protein and systolic blood pressure increased with rising rBMI in individuals with rBMI above zero, while these associations were minimal in individuals with rBMI below zero (Linear regression with hinge term, P=0.037 and P=0.026, respectively). In contrast, both the Matsuda index and body fat percentage showed steeper slopes when rBMI was below zero compared with above zero, with fat percentage increasing and Matsuda Index decreasing with rBMI (P=0.009 and P=1×10^-8^, respectively). These findings indicate that cardiometabolic markers do not respond uniformly across the rBMI spectrum. Inflammatory and blood pressure markers show stronger positive associations when BMI exceeds its genetically predicted value, whereas insulin sensitivity and body fat percentage show stronger associations when BMI is lower than genetically expected.

**Figure 3.**
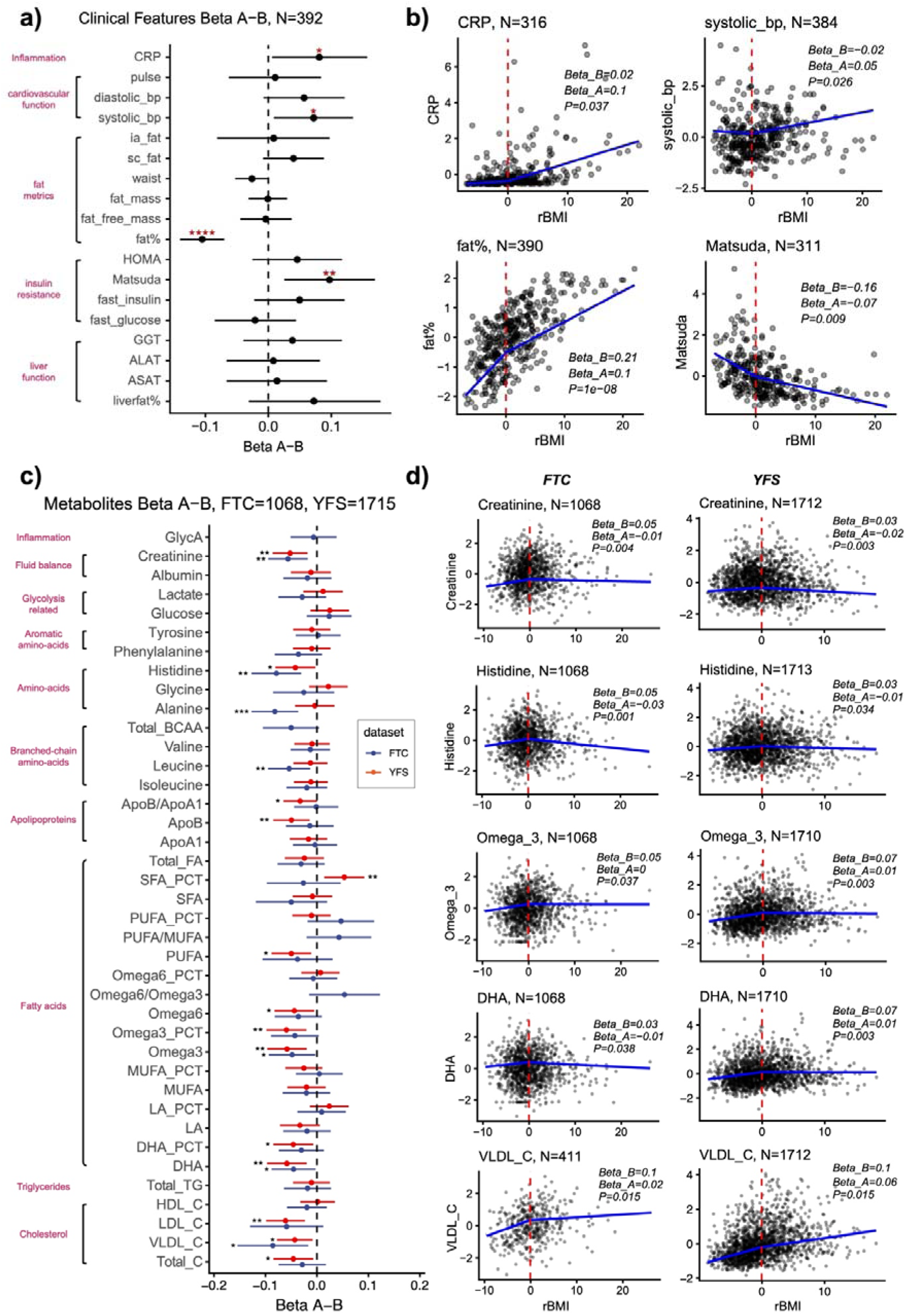
Differential effects of rBMI on clinical and metabolic biomarkers in FTC and YFS participants. **a)** Forest plots showing the coefficients (dots) with 95% confidence intervals (error bars) for the hinge term (Beta A-B) derived from linear mixed-effect models fitted in FTC participants. Clinical measurements (y-axis) were regressed on rBMI and the hinge variable, adjusting for age and sex as fixed effects and family ID as a random effect. **b)** Scatter plots showing the marginal effects of rBMI (x-axis) on clinical measurements (y-axis), for biomarkers exhibiting significantly different effect sizes below and above zero (hinge term P<0.05). Lines represent predictions from the fitted full model, showing expected outcomes across the rBMI range after correcting for age, sex and family ID. Regression coefficients below (Beta_B) and above (Beta_A) zero and P-value of the coefficient of hinge term were shown. **c)** the same as a), but for 39 clinically validated metabolic biomarkers as described by Julkunen et al. Blue indicates results from 1,068 FTC participants, and orange indicates results from 1,715 YFS participants. **d)** same as b), but for metabolic biomarkers significantly different in both FTC (left) and YFS (right). The number of points in each scatterplots are shown. Significance level: *, P<0.05; **, P<0.01; ***, P<0.001; ****, P<0.0001. Abbreviations: CRP, C-reactive protein; bp, blood pressure; ia, intra-abdominal; sc, subcutaneous; HOMA, the homeostatic model assessment used to quantify insulin resistance and beta-cell function; Matsuda, Matsuda index from oral glucose tolerance test; GGT, gamma-glutamyl transferase; ALAT, alanine transaminase; ASAT, aspartate aminotransferase; BCAA, branched-chain amino acid; SFA, saturated fatty acid; PUFA, polyunsaturated fatty acids; MUFA, monounsaturated fatty acids; LA, linoleic acid. _C, cholesterol.

#### Metabolomics

In metabolomic analyses from two independent cohorts (FTC, N=1,068 and YFS, N=1,715), five metabolites showed associations in the same direction in both cohorts with nominal significance (Linear regression with hinge term, P<0.05). Rising rBMI was associated with increasing levels of creatinine (P_FTC_=0.004, P_YFS_=0.003), histidine (P_FTC_=0.001, P_YFS_=0.034), omega-3 fatty acids (P_FTC_=0.037, P_YFS_=0.003), docosahexaenoic acid (DHA) (P_FTC_=0.038, P_YFS_=0.003), and VLDL cholesterol (P_FTC_=0.015, P_YFS_=0.015), when BMI was below the genetic prediction. The associations were negligible when BMI was above the genetic prediction (Fig. 3c). Scatterplots are shown in Fig. 3d.

#### DNA methylation and epigenetic aging

We next examined DNAm using 344,235 CpG sites common to the 450K and EPIC platforms. Meta-analyses were performed across four FTC sub-cohorts (OLD-EPIC, OLD-450K, YOUNG-EPIC, YOUNG-450K) and the YFS EPIC dataset (Methods). None of the CpG sites showed genome-wide significant association with above-versus below-prediction groups (P < 1×10^-7^) or remained significant after FDR correction (FDR<0.05). Six CpGs showed suggestive association (P < 1×10□□; Fig. 4a, Table S1), including cg09692695, cg11187796, cg10680180, cg19730092, cg21929875, and cg08595157. The six CpGs mapped to regulatory or gene body regions of biologically relevant genes, including *RTKN*, *ADRA2C*, *JAK2*, *ADAMTS14*, *VSIG2*, and *SCAPER* (Fig. 4b, Table S1).

**Figure 4.**
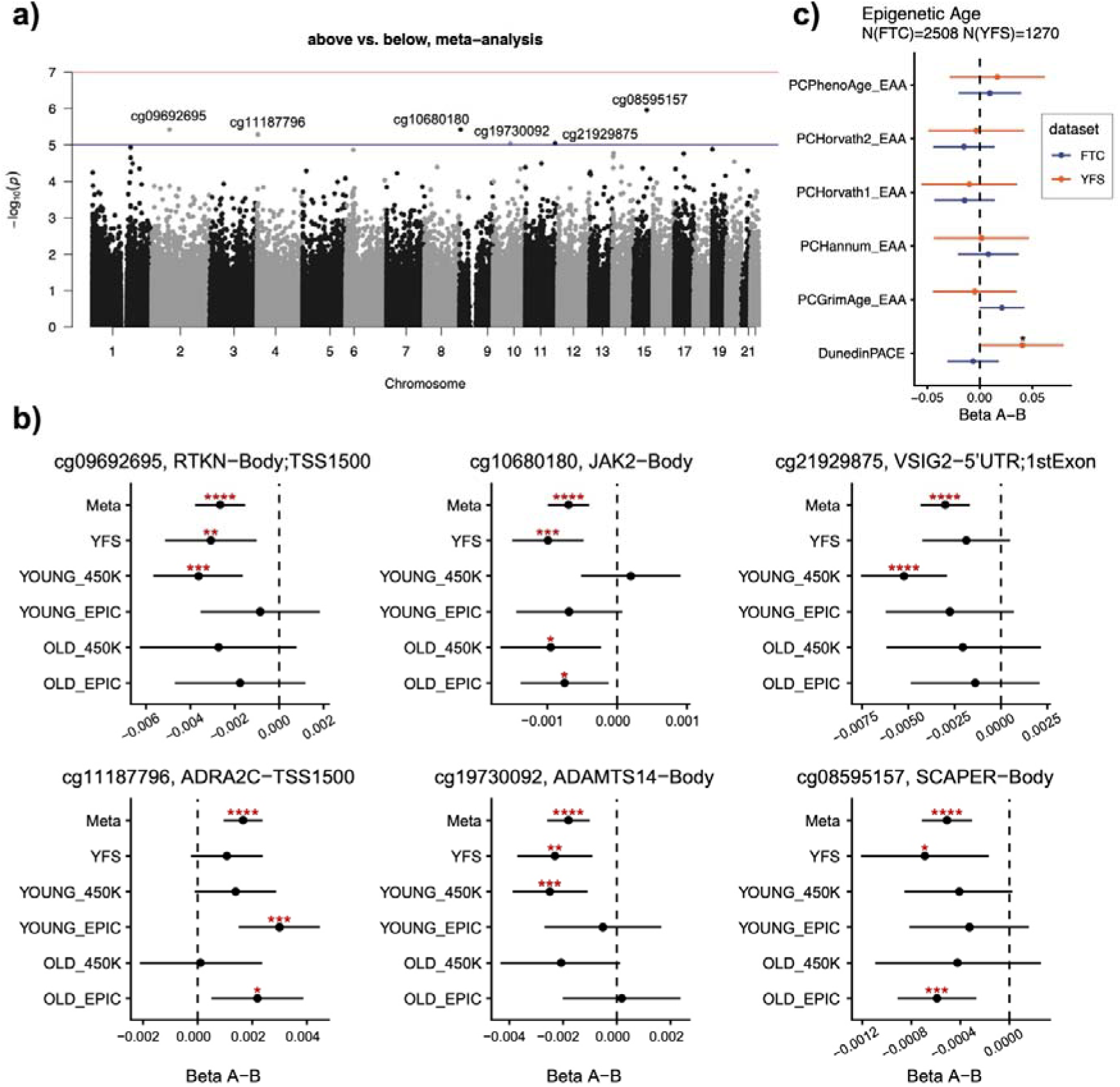
Differential effects of rBMI on DNAm in FTC and YFS participants. **a)** Manhattan plot showing the significance of associations for 347,521 CpG sites present in both 450K and EPIC platforms, based on a meta-analysis of five datasets. P-values correspond to the coefficients of the hinge term (Beta A-B) from linear mixed-effect regression models. Red line indicates the genome-wide significance threshold (P=1×10^-7^). Blue line indicates a suggestive significance threshold (P=1×10^-5^). CpGs with P<1×10^-5^ are annotated. **b)** Forest plots showing the coefficients of the hinge term (Beta A-B) for six CpG sites with P<1×10^-5^ in meta-analysis. Each panel corresponds to one CpG site and displays the effect estimate from the meta-analysis and the five individual datasets. Dots represent effect estimates and error bars indicate 95% confidence intervals. **c)** Forest plots showing the coefficients of the hinge term (Beta A-B) for epigenetic clocks. Blue indicates results from 2,508 FTC participants, and orange indicates results from 1,270 YFS participants. PCHorvath1, PC clock of Horvath’s pan-tissue clock. PCHorvath2, PC clock of Horvath’s skin and blood clock. Significance level: *, P<0.05; **, P<0.01; ***, P<0.001; ****, P<0.0001.

Across all epigenetic clocks analyzed in FTC and most of them in YFS, rising rBMI was associated with increasing epigenetic age acceleration (EAA) (Fig. S4). The magnitude of EAA was largely independent of whether BMI was below or above the genetic prediction (Linear regression, P < 0.05; Fig. 4c). The only exception was DunedinPACE in YFS, which showed a marginally steeper increase when BMI exceeded genetic prediction (P=0.04). Overall, these results suggest that EAA is related to excess BMI *per se* rather than to the deviation from the genetic prediction of BMI.

#### Proteomics

To explore protein-level effects, we analyzed plasma proteomic data from 381 FTC individuals for 2,321 proteins. Although no proteins reached the FDR < 0.05 threshold, 342 proteins exhibited nominally significant differences in their associations with rBMI when BMI is above or below genetic prediction, several of which have known links to obesity (Fig. 5a). Among these, leptin (LEP) showed the strongest increase with rising rBMI, with a markedly steeper slope when BMI was below the genetic prediction (Fig. 5b). KEGG pathway enrichment analysis of these proteins revealed overrepresentation of cytokine-cytokine receptor interaction, PI3K-Akt signaling, immunoglobulin superfamily cell adhesion molecule signaling, cell adhesion molecule interaction, and JAK-STAT signaling pathways (Fig. 5c).

**Figure 5.**
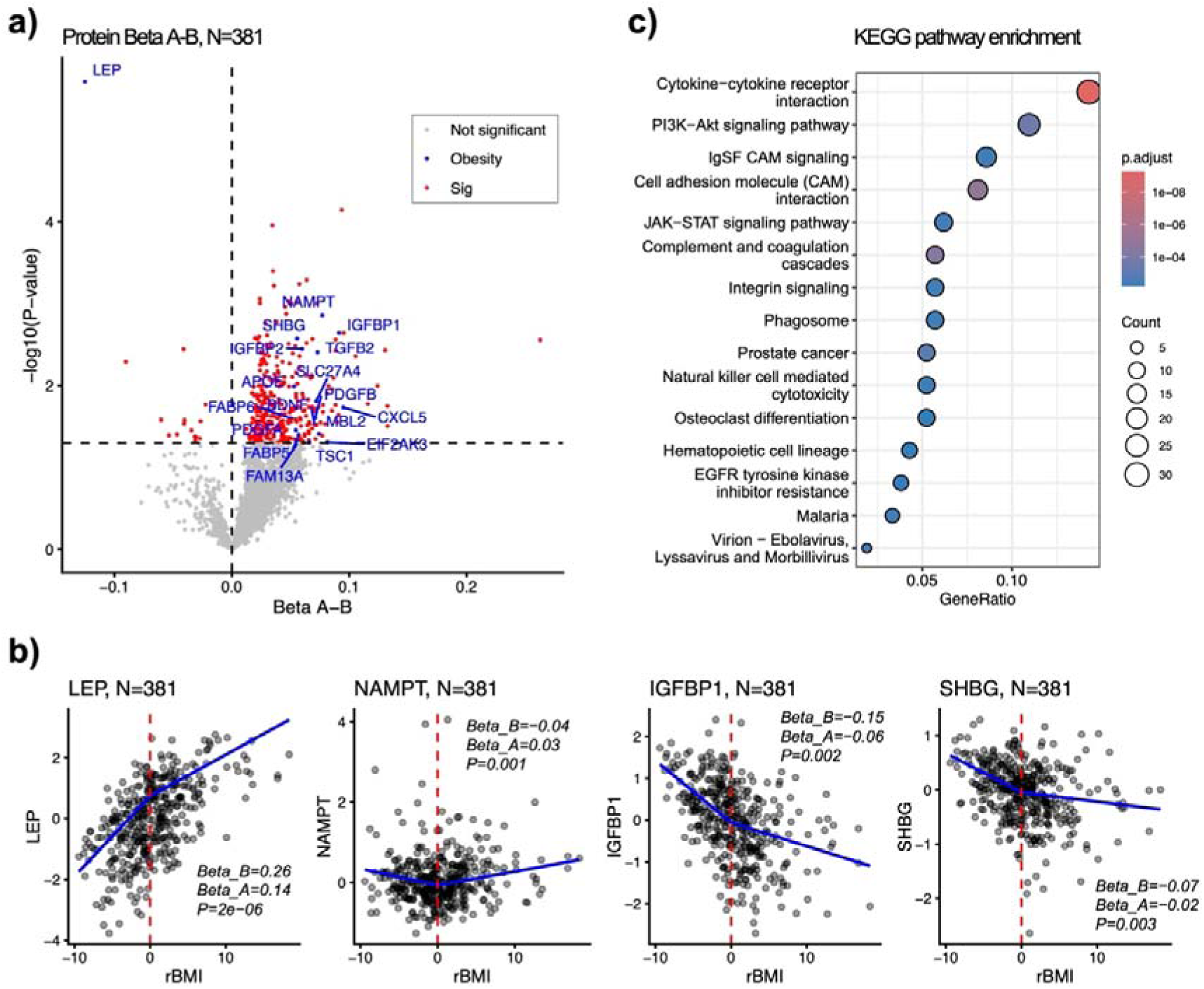
Differential effects of rBMI on proteome in FTC participants. **a)** Volcano plot showing the -log_10_(P) values (y-axis) and regression coefficients of the hinge term (x-axis) for 2,321 analyzed proteins. Proteins exhibiting significantly different expression in above vs. below zero were highlighted in red; among those, proteins with known obesity-related functions are annotated in blue. **b)** Scatter plots showing the marginal effects of rBMI (x-axis) on protein abundance (y-axis) for the proteins with a significant hinge term (P<0.05). Lines represent predictions from the fitted full model, showing expected outcomes across the rBMI range after correcting for age, sex and family ID. Regression coefficients below (Beta_B) and above (Beta_A) zero, as well as the P-value for the hinge term, are shown. **c)** Dot plot showing the top 15 enriched KEGG pathways among 342 proteins with P<0.05. The x-axis indicates the gene ratio, circle size represents the number of proteins in each pathway, and the color indicates the enrichment P-values.

Overall, the molecular level findings indicate that when BMI is below the genetic prediction, metabolites, protein profiles, and several signaling pathways respond more sensitively to BMI, while individuals whose BMI is above genetic expectation show weaker or negligible molecular changes across these layers, with EAA reflecting BMI itself rather than deviation from its genetic prediction.

### Within-pair analyses of BMI-discordant twin pairs in FTC

We next evaluated these associations in BMI-discordant twin pairs grouped according to alignment with genetically predicted BMI (Group A, higher-BMI being above prediction i.e. lower-BMI following the genetic prediction; Group B, lower-BMI being below prediction i.e. higher-BMI following the genetic prediction) (Methods). We examined whether within-pair differences in rBMI were associated with differences in clinical, epigenetic, metabolomic, and proteomic traits, and whether these associations differed between the groups.

#### Clinical measurements

Systolic blood pressure and gamma-glutamyl transferase increased in the higher-BMI twins in both groups, and they increased more steeply with rBMI in Group A compared with Group B (linear regression, P = 0.004 and P = 0.03, respectively) (Fig. 6a–b). None of the associations showed statistically significant group-specific effects after multiple-testing correction (FDR < 0.05).

**Figure 6.**
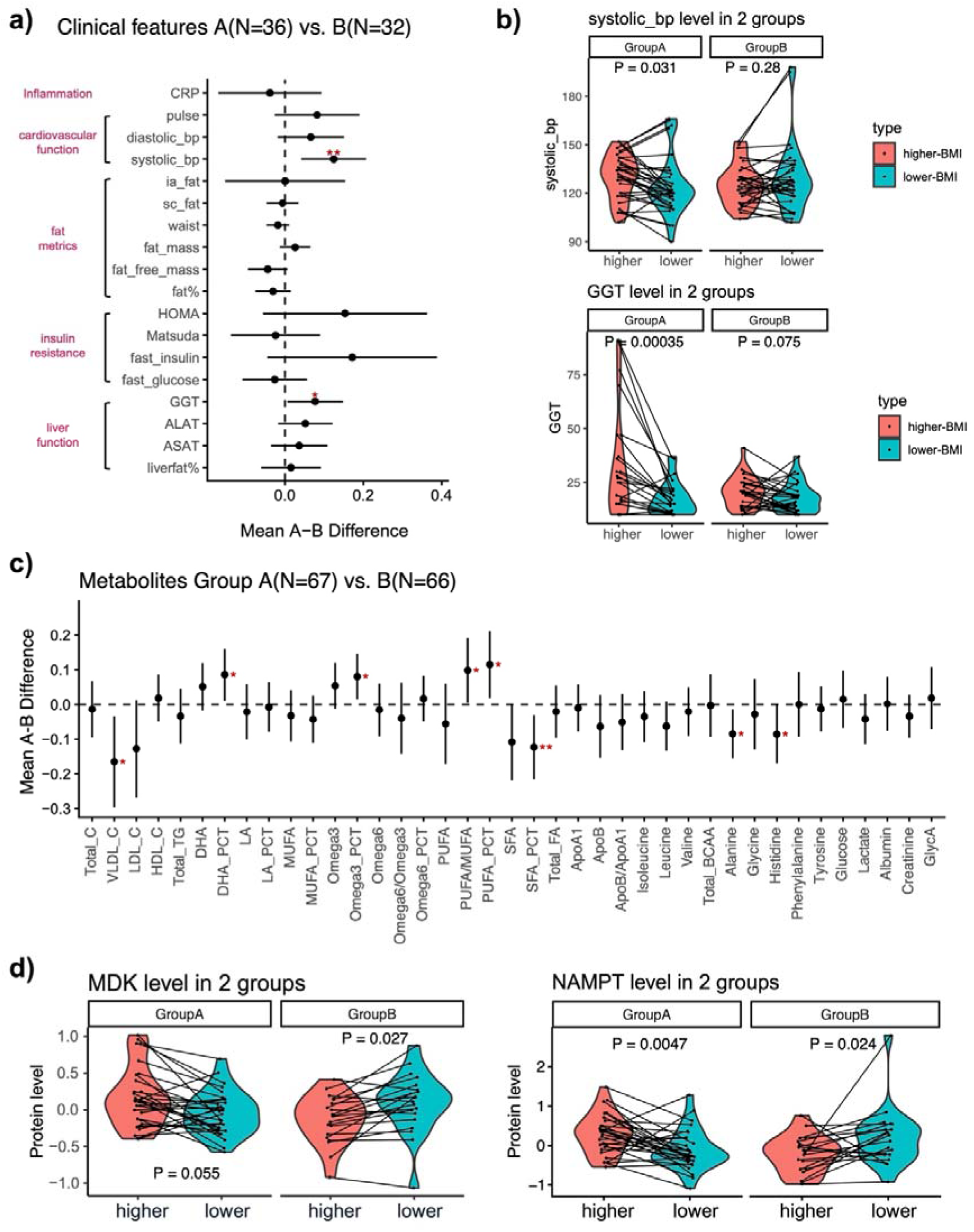
Group-specific clinical and omics measurements in BMI-discordant twin pairs. **a)** Dot plot showing the coefficients (dots) and 95% confidence intervals (error bars) derived from fixed-effect within-pair linear regression using twin pairs from the two groups, where clinical measurements (y-axis) were regressed against rBMI interacting with the group variable. **b)** Up: Violin plot showing the actual systolic blood pressure level in two groups, colored by higher-vs. lower BMI co-twins (red indicates higher-BMI co-twins, blue indicates lower-BMI co-twins). Down: Same plot for blood gamma-glutamyl transferase (GGT) levels. Each line connects individuals in a twin pair. P-value is from paired Wilcoxon Test. **c)** Same as a), but for 39 metabolic markers. **d)** Violin plot showing normalized MDK and NAMPT protein level (y-axis) in two groups, colored by higher vs. lower-BMI co-twins (red indicates higher-BMI co-twins, blue indicates lower-BMI co-twins). P-values were derived from paired Wilcoxon Rank Sum Test. Significance level: *, P<0.05; **, P<0.01; ***, P<0.001; ****, P<0.0001. Group A: BMI-discordant (delta BMI>3) same-sex twin pairs with the higher-BMI co-twin’s BMI above genetic prediction and the lower-BMI co-twin’s BMI within genetic prediction; Group B: BMI-discordant (delta BMI>3) same-sex twin pairs with the higher-BMI co-twin’s BMI within genetic prediction and the lower-BMI co-twin’s BMI below genetic prediction.

#### Metabolomics

In metabolomic analyses, several metabolites showed group-specific associations at the nominal P-value level (Fig. 6c). Very-low-density lipoprotein cholesterol (VLDL-C), percentage of saturated fatty acids (SFA), alanine, histidine, DHA and omega-3 fatty acids were positively associated with rBMI in both groups, whereas polyunsaturated fatty acids (PUFA) showed a negative association (Fig. S5). Although the directions of these associations were consistent across groups, their effect sizes differed. VLDL-C (P = 0.01 for difference between groups), percentage of SFA (P = 0.01), alanine (P = 0.02), histidine (P = 0.05) and PUFA (P=0.02) were more strongly associated with rBMI in Group B compared to Group A. In contrast, DHA (P = 0.02) and omega-3 fatty acids (P = 0.02) showed stronger associations with rBMI in Group A (Fig. 6c, S5). None of the differences between the groups remained significant after FDR correction (FDR < 0.05).

#### DNA methylation and epigenetic aging

DNA methylation analyses revealed no group-specific differences in individual CpG sites after correction for multiple testing (FDR < 0.05). Two CpGs showed suggestive association (P < 1×10□□), including cg26558023 and cg01034631. Both CpGs are located in open sea regions with limited functional annotation. cg01034631 maps to the lncRNA *MEG8* within the imprinted DLK1-DIO3 locus, although its relevance to BMI remains unclear. Across all epigenetic clocks, no group-specific effect was observed (Fig. S6).

#### Proteomics

In within-pair proteomic analyses, several proteins showed nominal group-specific associations with rBMI. Among top nominally significant proteins, MDK and NAMPT displayed opposite directional associations between groups, increasing in the higher-BMI co-twins in Group A but decreasing in Group B (Fig. 6d). SERPINA7 showed a stronger positive association in Group A, increasing in the higher-BMI co-twins, whereas LDLR showed a stronger positive association in Group B (Fig. S7). None remained significant after multiple testing correction.

Overall, the within-pair analyses in two groups showed a pattern consistent with the hinge regression results, largely characterized by no differences between groups or stronger associations in the Group B where the higher-BMI co-twin aligned with their genetically predicted BMI.

## Discussion

In this study, we quantified deviation from genetic predisposition to BMI using a PRS-informed residual BMI measure (rBMI) and explored its molecular correlates across multiple biological layers in two Finnish population cohorts. We found the associations were generally stronger when BMI was below the genetically predicted BMI, while they were weaker or negligible when BMI was above the genetic prediction across the clinical and multi-omics traits. Considering how BMI aligns with genetic predisposition may provide a more refined framework for understanding variability in metabolic health and disease risk.

Importantly, the observed differences in slope should be interpreted in the context of the underlying BMI distribution: individuals exceeding their genetic prediction can reach substantially higher BMI values, whereas those below prediction are constrained by a lower physiological bound. As such, steeper associations below genetic prediction may partly reflect this restricted range, whereas steeper associations above genetic prediction are less likely to be explained by scale alone and may instead indicate a more pronounced or potentially pathological relationship. In this context, the stronger positive associations of C-reactive protein and systolic blood pressure above genetic prediction suggest a greater systemic inflammation and vascular burden in individuals whose BMI exceeds their genetic predisposition. Meanwhile, the steeper associations of Matsuda index and body fat percentage below genetic prediction suggest that these traits are more sensitive to BMI variation at lower levels of adiposity. Together, these observations highlight that the physiological consequences of BMI are not uniform but depend on its deviation from genetic expectation, supporting the idea that rBMI provides additional insight into metabolic health beyond absolute BMI alone.

In the DNAm analysis, the top CpGs mapped to genes involved in leptin signaling (*JAK2*) (35), neuroendocrine regulation (*ADRA2C*) (36), adiposity (*ADAMTS14*) (37), cellular stress (*RTKN*) (38), cell-cycle progression (*SCAPER*) (39) and cardiovascular risk (*VSIG2*) (40). However, the very small effect sizes (∼0.001) indicate that these associations should be interpreted with caution. Epigenetic age acceleration has been shown to robustly associate with BMI (17), and here we further show that EAA is associated with BMI irrespective of its deviation from the genetically predicted BMI, implying that EAA reflects cumulative weight gain regardless of the degree of genetic mismatch. In proteomics analysis, leptin (LEP) increased with rBMI and the positive association was stronger when BMI was below genetic prediction. This pattern is consistent with leptin resistance at higher adiposity levels, where additional weight gain leads to weaker leptin signaling despite increased fat mass (41). Protein pathway enrichment highlighted cytokine-cytokine receptor interaction, PI3K-Akt, and JAK-STAT pathways, suggesting coordinated changes in inflammatory and metabolic signaling in response to deviation from genetically predicted BMI.

We further conducted within-pair analyses in BMI-discordant twin pairs grouped by alignment with genetic prediction. This design allowed us to go beyond the partial genetic information captured by PRS and to control for the genetics and environment shared within each twin pair. Several within-pair differences were consistent with the population-level analysis. Specifically, systolic blood pressure and gamma-glutamyl transferase (GGT) showed stronger positive associations with rBMI in pairs where the higher-BMI twin exceeded their genetic expectation (Group A), consistent with greater physiological strain beyond genetically predicted BMI. In contrast, metabolomic patterns in pairs where the higher-BMI twin aligned with genetic prediction (Group B) had relatively larger effect sizes in VLDL cholesterol, saturated fatty acids, alanine, histidine, and PUFA compared with pairs where the lower-BMI twin followed the genetic prediction, possibly reflecting early metabolic adaptation before reaching a pathological threshold (42). rBMI was generally associated with similar epigenetic aging and proteomic responses across the groups. Notably, Midkine (MDK), an adipocyte-secreted growth factor associated with obesity and insulin resistance (43), as well as NAMPT, likely reflecting its extracellular form (eNAMPT), an adipokine with elevated circulating levels in obesity (44), showed divergent associations between the groups, with the lowest levels near genetically expected BMI. The lowest levels of both MDK and NAMPT near genetically predicted BMI further support the notion that alignment of genetic predisposition and observed BMI represents a more metabolically stable state, whereas deviation in either direction may be accompanied by dysregulated adipocyte signaling. Together, the convergence between population-level and within-pair analyses supports the presence of subtle but consistent multi-omic patterns to deviations from genetically determined limits of adiposity.

A key strength of this study is the integration of population-level and genetically controlled twin analyses across multiple omics layers. The population-level hinge models provided sensitivity to detect differential patterns, while the twin-pair analyses offered specificity by controlling for shared genetics and early-life environment. Importantly, in unrelated individuals, deviation from PRS-predicted BMI may still partly reflect unmeasured genetic effects, as current PRS capture only a fraction of heritable variation. While such deviations are often interpreted as environmentally driven, twin designs provide stronger control for genetic background—complete in monozygotic pairs and 50% on average in dizygotic pairs. Although both MZ and DZ pairs were included due to the limited power of MZ pairs alone, our approach still controls for more than 50% of genetic variation. Notably, the consistency between population-level and within-pair analyses suggests that variation in rBMI is likely driven to a substantial extent by non-genetic factors. In this context, grouping twins based on deviation from PRS-predicted BMI represents a novel framework that extends beyond traditional high-versus low-PRS stratification and avoids arbitrary risk thresholds that may vary across populations and PRS construction methods.

Several limitations should be acknowledged. First, despite being one of the largest twin cohorts with multi-omics data, statistical power may have been limited for within-pair analysis due to small sample sizes. However, considering twins share a large percentage of their segregating genome and are raised together in the same family environment, BMI-discordant twin pairs are rare in the first place. Second, rBMI remains correlated with observed BMI, as current PRS explain only ∼5% of BMI variance in our cohort (8.4% in the discovery GWAS (5), 17.6% in the newest GWAS (6)). Thus, we emphasize that rBMI is not intended to be independent of BMI, but rather to reparameterize BMI relative to genetic predisposition. Consequently, observed associations with rBMI may partly reflect BMI itself, although rBMI provides additional insight into whether biological processes differ depending on deviation from genetic expectation. Additionally, while significantly correlated with a phenotype at the population level, different PRSes can give highly variable individual-level estimates of risk (45,46). Improving the predictive accuracy of PRS for BMI could significantly enhance our understanding of this complex trait. For instance, future studies could leverage whole-genome sequencing data, which can explain up to 30% SNP heritability in BMI (47,48), to construct more comprehensive PRS models. Finally, the underlying biological mechanisms accounting for rBMI remain to be determined and require targeted experimental studies and validation in independent samples.

In summary, our findings demonstrate that deviations from genetically predicted BMI capture subtle yet consistent molecular and clinical signatures linked to adiposity and metabolic health. Looking ahead, incorporating longitudinal data and detailed environmental measures could help explain how genetic and environmental factors interact over time, and whether identified associations are likely causal. Future work could also explore PRSes based on traits more closely related to metabolically adverse adiposity, such as visceral adipose tissue or anthropometric proxies like waist-to-height ratio, which may better capture cardiometabolic risk than BMI. Finally, a key priority is to explore whether rBMI and associated multi-omics signatures correlate with meaningful health outcomes, such as cardiovascular diseases and liver dysfunction. Comprehensively evaluating these relationships across diverse populations will be an important next step in this line of research.

## Conclusion

Deviations from genetically predicted BMI were associated with distinct clinical and multi-omics profiles, with differences depending on whether BMI was above or below genetic expectation. Integrating deviation from genetic predisposition may improve characterizing metabolic heterogeneity and identify individuals at higher cardiometabolic risk.

## Acknowledgements

The authors thank all the study participants from the Finnish Twin Cohort and Young Finns Study for their long-term contribution. We also acknowledge the computational resources and technical support provided by the Institute for Molecular Medicine Finland (FIMM) Technology Center, University of Helsinki IT center, and CSC – IT Center for Science, Finland. We thank Dr. Jadwiga Buchwald for her advice in statistical modeling, and Dr. Birgitta van der Kolk and Dr. Maheswary Muniandy for their contribution to the generation of data for the FTC metabolic sub-cohort data and for their helpful comments on the manuscript. Figure 1a was created with <u>BioRender.com</u>. The authors used ChatGPT5.2 (OpenAI) to assist with language polishing of the manuscript. All content was reviewed and verified by the authors.

## Ethical approval and consent to participant

The data collection of FTC has been approved by the appropriate national research ethics committees, with the most recent approval granted by the Hospital District of Helsinki and Uusimaa ethics board in 2018 (#1799/2017). Ethical approval for the metabolic substudy was obtained from the Ethical Review Board of Helsinki University Hospital (#270/13/03/01/2008, 6.8.08, 3.4.13, 16.3.2016). The data collection of YFS was approved by the Ethics Committee of the Hospital District of Southwest Finland on June 20, 2017 (ETMK: 68/1801/2017). All participants gave their written informed consent, and the studies were conducted in accordance with the Declaration of Helsinki.

## Data availability

FTC: The phenotype and omics data from the Finnish Twin Cohort used in this study are stored in the Biobank of the Finnish Institute for Health and Welfare (Helsinki, Finland). Qualified researchers may obtain these data through a standardized application procedure (https://thl.fi/en/statistics-and-data/data-and-services/research-use-and-data-permits).

YFS: The dataset supporting the conclusions of this article were obtained from the Cardiovascular Risk In Young Finns study which comprises health related participant data. The use of data is restricted under the regulations on professional secrecy (Act on the Openness of Government Activities, 612/1999) and on sensitive personal data (Personal Data Act, 523/1999, implementing the EU data protection directive 95/46/EC). Due to these restrictions, the data can not be stored in public repositories or otherwise made publicly available. Data access may be permitted on a case by case basis upon request only. Data sharing outside the group is done in collaboration with YFS group and requires a data-sharing agreement. Investigators can submit an expression of interest to the chairman of the publication committee (Prof Mika Kähönen, Tampere University, Finland).

## Author Contributions

MO conceived the study. TZ performed the main analyses and drafted the manuscript. MO and RC co-supervised this study and provided critical feedback. AH and MH preprocessed the FTC DNAm data and computed epigenetic age scores. GD preprocessed the FTC metabolomics and proteomics data. TP processed the FTC genotype data and calculated the PRS for BMI. BJB processed longitudinal BMI information of FTC participants and provided analytical feedback. KHP provided clinical data for the FTC metabolic subcohort. JK, principal investigator of the FTC, co-designed phenotype and DNA collection and contributed to the statistical analysis strategy. ER, OR, TL and MAK provided the YFS phenotypes, DNAm and metabolomics data. LPL calculated the PRS for BMI for YFS participants. All authors discussed the results, contributed to manuscript revisions, and approved the final version of the manuscript.

## Supplementary Information

Additional file 1: .docx, Supplementary Figures S1-7 and Supplementary Methods.

Additional file 2: .xlsx, Supplementary Table S1-2 and statistical results for the main figures.

## References

1. Elks CE, Den Hoed M, Zhao JH, Sharp SJ, Wareham NJ, Loos RJF, et al. Variability in the Heritability of Body Mass Index: A Systematic Review and Meta-Regression. Front Endocrinol. 2012;3. doi:10.3389/fendo.2012.00029

2. Maes HHM, Neale MC, Eaves LJ. Genetic and environmental factors in relative body weight and human adiposity. Behav Genet. 1997;27(4):325–51. doi:10.1023/A:1025635913927

3. Drouard G, Hagenbeek FA, Whipp AM, Pool R, Hottenga JJ, Jansen R, et al. Longitudinal multi-omics study reveals common etiology underlying association between plasma proteome and BMI trajectories in adolescent and young adult twins. BMC Med. 2023 Dec 21;21(1):508. doi:10.1186/s12916-023-03198-7

4. Silventoinen K, Jelenkovic A, Sund R, Yokoyama Y, Hur YM, Cozen W, et al. Differences in genetic and environmental variation in adult BMI by sex, age, time period, and region: an individual-based pooled analysis of 40 twin cohorts. Am J Clin Nutr. 2017 Aug;106(2):457–66. doi:10.3945/ajcn.117.153643

5. Yengo L, Sidorenko J, Kemper KE, Zheng Z, Wood AR, Weedon MN, et al. Meta-analysis of genome-wide association studies for height and body mass index in ∼700000 individuals of European ancestry. Hum Mol Genet. 2018 Oct 15;27(20):3641–9. doi:10.1093/hmg/ddy271

6. Smit RAJ, Wade KH, Hui Q, Arias JD, Yin X, Christiansen MR, et al. Polygenic prediction of body mass index and obesity through the life course and across ancestries. Nat Med. 2025 Jul 21. doi:10.1038/s41591-025-03827-z

7. Ojalehto E, Zhan Y, Jylhävä J, Reynolds CA, Dahl Aslan AK, Karlsson IK. Genetically and environmentally predicted obesity in relation to cardiovascular disease: a nationwide cohort study. eClinicalMedicine. 2023 Apr;58:101943. doi:10.1016/j.eclinm.2023.101943

8. Zilhao NR, Zhang J, Boomsma DI, Sørensen TIA, Dahm CC. Deviation From Genetically Predicted BMI and All-Cause Mortality: A Cohort Study in the UK Biobank. Obesity. 2025 Oct 2;oby.70042. doi:10.1002/oby.70042

9. Geurts YM, Dugué PA, Joo JE, Makalic E, Jung CH, Guan W, et al. Novel associations between blood DNA methylation and body mass index in middle-aged and older adults. Int J Obes. 2018 Apr;42(4):887–96. doi:10.1038/ijo.2017.269

10. Wahl S, Drong A, Lehne B, Loh M, Scott WR, Kunze S, et al. Epigenome-wide association study of body mass index, and the adverse outcomes of adiposity. Nature. 2017 Jan;541(7635):81–6. doi:10.1038/nature20784

11. MacTel Consortium, Lotta LA, Pietzner M, Stewart ID, Wittemans LBL, Li C, et al. A cross-platform approach identifies genetic regulators of human metabolism and health. Nat Genet. 2021 Jan;53(1):54–64. doi:10.1038/s41588-020-00751-5

12. Rodriguez-Muñoz A, Motahari-Rad H, Martin-Chaves L, Benitez-Porres J, Rodriguez-Capitan J, Gonzalez-Jimenez A, et al. A Systematic Review of Proteomics in Obesity: Unpacking the Molecular Puzzle. Curr Obes Rep. 2024 May 4;13(3):403–38. doi:10.1007/s13679-024-00561-4

13. Drouard G, Argentieri MA, Heikkinen A, Ollikainen M, Kaprio J. Associations Between 40-Year Trajectories of BMI and Proteomic and Epigenetic Aging Clocks: Deciphering Nonlinearity and Interactions. Aging Cell. 2026 Feb;25(2):e70397. doi:10.1111/acel.70397

14. Föhr T, Hendrix A, Kankaanpää A, Laakkonen EK, Kujala U, Pietiläinen KH, et al. Metabolic syndrome and epigenetic aging: a twin study. Int J Obes. 2024 Jun;48(6):778–87. doi:10.1038/s41366-024-01466-x

15. Higgins-Chen AT, Thrush KL, Wang Y, Minteer CJ, Kuo PL, Wang M, et al. A computational solution for bolstering reliability of epigenetic clocks: implications for clinical trials and longitudinal tracking. Nat Aging. 2022 Jul 15;2(7):644–61. doi:10.1038/s43587-022-00248-2

16. Levine ME, Lu AT, Quach A, Chen BH, Assimes TL, Bandinelli S, et al. An epigenetic biomarker of aging for lifespan and healthspan. Aging. 2018 Apr 18;10(4):573–91. doi:10.18632/aging.101414

17. Lundgren S, Kuitunen S, Pietiläinen KH, Hurme M, Kähönen M, Männistö S, et al. BMI is positively associated with accelerated epigenetic aging in twin pairs discordant for body mass index. J Intern Med. 2022 Oct;292(4):627–40. doi:10.1111/joim.13528

18. Oh HSH, Rutledge J, Nachun D, Pálovics R, Abiose O, Moran-Losada P, et al. Organ aging signatures in the plasma proteome track health and disease. Nature. 2023 Dec 7;624(7990):164–72. doi:10.1038/s41586-023-06802-1

19. Naukkarinen J, Rissanen A, Kaprio J, Pietiläinen KH. Causes and consequences of obesity: the contribution of recent twin studies. Int J Obes. 2012 Aug;36(8):1017–24. doi:10.1038/ijo.2011.192

20. Kaidesoja M, Aaltonen S, Bogl LH, Heikkilä K, Kaartinen S, Kujala UM, et al. FinnTwin16: A Longitudinal Study from Age 16 of a Population-Based Finnish Twin Cohort. Twin Res Hum Genet. 2019 Dec;22(6):530–9. doi:10.1017/thg.2019.106

21. Kaprio J, Bollepalli S, Buchwald J, Iso-Markku P, Korhonen T, Kovanen V, et al. The Older Finnish Twin Cohort — 45 Years of Follow-up. Twin Res Hum Genet. 2019 Aug;22(4):240–54. doi:10.1017/thg.2019.54

22. Rose RJ, Salvatore JE, Aaltonen S, Barr PB, Bogl LH, Byers HA, et al. FinnTwin12 Cohort: An Updated Review. Twin Res Hum Genet. 2019 Oct;22(5):302–11. doi:10.1017/thg.2019.83

23. Åkerblom HK, Uhari M, Pesonen E, Dahl M, Kaprio EA, Nuutinen EM, et al. Cardiovascular Risk in Young Finns. Ann Med. 1991 Jan;23(1):35–9. doi:10.3109/07853899109147928

24. Pahkala K, Rovio S, Kartiosuo N, Auranen K, Bourgery M, Elovainio M, et al. Cohort Profile Update: Expanding the Cardiovascular Risk in Young Finns Study into a multigenerational cohort. Int J Epidemiol. 2026 Jan 2;55(1):dyaf206. doi:10.1093/ije/dyaf206

25. Raitakari OT, Juonala M, Ronnemaa T, Keltikangas-Jarvinen L, Rasanen L, Pietikainen M, et al. Cohort Profile: The Cardiovascular Risk in Young Finns Study. Int J Epidemiol. 2008 Dec 1;37(6):1220–6. doi:10.1093/ije/dym225

26. Berntzen BJ, Palviainen T, Silventoinen K, Pietiläinen KH, Kaprio J. Polygenic risk of obesity and BMI trajectories over 36 years: A longitudinal study of adult Finnish twins. Obesity. 2023 Dec;31(12):3086–94. doi:10.1002/oby.23906

27. Tuomela J, Kaprio J, Sipilä PN, Silventoinen K, Wang X, Ollikainen M, et al. Accuracy of self-reported anthropometric measures — Findings from the Finnish Twin Study. Obes Res Clin Pract. 2019 Nov;13(6):522–8. doi:10.1016/j.orcp.2019.10.006

28. Van Der Kolk BW, Saari S, Lovric A, Arif M, Alvarez M, Ko A, et al. Molecular pathways behind acquired obesity: Adipose tissue and skeletal muscle multiomics in monozygotic twin pairs discordant for BMI. Cell Rep Med. 2021 Apr;2(4):100226. doi:10.1016/j.xcrm.2021.100226

29. Soininen P, Kangas AJ, Würtz P, Tukiainen T, Tynkkynen T, Laatikainen R, et al. High-throughput serum NMR metabonomics for cost-effective holistic studies on systemic metabolism. The Analyst. 2009;134(9):1781. doi:10.1039/b910205a

30. Soininen P, Kangas AJ, Würtz P, Suna T, Ala-Korpela M. Quantitative Serum Nuclear Magnetic Resonance Metabolomics in Cardiovascular Epidemiology and Genetics. Circ Cardiovasc Genet. 2015 Feb;8(1):192–206. doi:10.1161/CIRCGENETICS.114.000216

31. Masip G, Silventoinen K, Keski-Rahkonen A, Palviainen T, Sipilä PN, Kaprio J, et al. The genetic architecture of the association between eating behaviors and obesity: combining genetic twin modeling and polygenic risk scores. Am J Clin Nutr. 2020 Oct;112(4):956–66. doi:10.1093/ajcn/nqaa181

32. Borodulin K, Tolonen H, Jousilahti P, Jula A, Juolevi A, Koskinen S, et al. Cohort Profile: The National FINRISK Study. Int J Epidemiol. 2018 Jun 1;47(3):696–696i. doi:10.1093/ije/dyx239

33. Gunasekara FI, Richardson K, Carter K, Blakely T. Fixed effects analysis of repeated measures data. Int J Epidemiol. 2014 Feb;43(1):264–9. doi:10.1093/ije/dyt221

34. Rose RJ, Latvala A, Silventoinen K, Kaprio J. Alcohol consumption at age 18–25 and number of children at a 33-year follow-up: Individual and within-pair analyses of Finnish twins. Alcohol Clin Exp Res. 2022 Aug;46(8):1552–64. doi:10.1111/acer.14886

35. Dodington DW, Desai HR, Woo M. JAK/STAT – Emerging Players in Metabolism. Trends Endocrinol Metab. 2018 Jan;29(1):55–65. doi:10.1016/j.tem.2017.11.001

36. Braun K, Oeckl J, Westermeier J, Li Y, Klingenspor M. Non-adrenergic control of lipolysis and thermogenesis in adipose tissues. Suarez RK, Hoppeler HH, editors. J Exp Biol. 2018 Mar 7;221(Suppl_1):jeb165381. doi:10.1242/jeb.165381

37. Kentistou KA, Luan J, Wittemans LBL, Hambly C, Klaric L, Kutalik Z, et al. Large scale phenotype imputation and in vivo functional validation implicate ADAMTS14 as an adiposity gene. Nat Commun. 2023 Jan 19;14(1):307. doi:10.1038/s41467-022-35563-0

38. Liu CA, Wang MJ, Chi CW, Wu CW, Chen JY. Rho/Rhotekin-mediated NF-κB activation confers resistance to apoptosis. Oncogene. 2004 Nov 18;23(54):8731–42. doi:10.1038/sj.onc.1208106

39. Tsang WY, Wang L, Chen Z, Sánchez I, Dynlacht BD. SCAPER, a novel cyclin A–interacting protein that regulates cell cycle progression. J Cell Biol. 2007 Aug 13;178(4):621–33. doi:10.1083/jcb.200701166

40. Plaza-Florido A, Olvera-Rojas M, Alcantara JMA, Radom-Aizik S, Ortega FB. Targeted proteomics involved in cardiovascular health and heart rate variability in children with overweight/obesity. Am J Hum Biol. 2024 Sep;36(9):e24113. doi:10.1002/ajhb.24113

41. Ladyman SR, Grattan DR. JAK-STAT and feeding. JAK-STAT. 2013 Apr;2(2):e23675. doi:10.4161/jkst.23675

42. Laclaustra M, Lopez-Garcia E, Civeira F, Garcia-Esquinas E, Graciani A, Guallar-Castillon P, et al. LDL Cholesterol Rises With BMI Only in Lean Individuals: Cross-sectional U.S. and Spanish Representative Data. Diabetes Care. 2018 Oct 1;41(10):2195–201. doi:10.2337/dc18-0372

43. Fan N, Sun H, Wang Y, Zhang L, Xia Z, Peng L, et al. Midkine, a Potential Link between Obesity and Insulin Resistance. Lluch GL, editor. PLoS ONE. 2014 Feb 7;9(2):e88299. doi:10.1371/journal.pone.0088299

44. Romacho T, Valencia I, Ramos-González M, Vallejo S, López-Esteban M, Lorenzo O, et al. Visfatin/eNampt induces endothelial dysfunction in vivo: a role for Toll-Like Receptor 4 and NLRP3 inflammasome. Sci Rep. 2020 Mar 25;10(1):5386. doi:10.1038/s41598-020-62190-w

45. Abramowitz SA, Boulier K, Keat K, Cardone KM, Shivakumar M, DePaolo J, et al. Evaluating Performance and Agreement of Coronary Heart Disease Polygenic Risk Scores. JAMA. 2025 Jan 7;333(1):60. doi:10.1001/jama.2024.23784

46. Loos RJF, Yeo GSH. The genetics of obesity: from discovery to biology. Nat Rev Genet. 2022 Feb;23(2):120–33. doi:10.1038/s41576-021-00414-z

47. Wainschtein P, Jain D, Zheng Z, TOPMed Anthropometry Working Group, Aslibekyan S, Becker D, et al. Assessing the contribution of rare variants to complex trait heritability from whole-genome sequence data. Nat Genet. 2022 Mar;54(3):263–73. doi:10.1038/s41588-021-00997-7

48. Wainschtein P, Zhang Y, Schwartzentruber J, Kassam I, Sidorenko J, Fiziev PP, et al. Estimation and mapping of the missing heritability of human phenotypes. Nature. 2025 Nov 12. doi:10.1038/s41586-025-09720-6

