## Additional file 1 for "BMI Deviations from Genetic Prediction Are Associated with Distinct Molecular Signatures in Finnish Population Cohorts"

**Supplementary Figures**

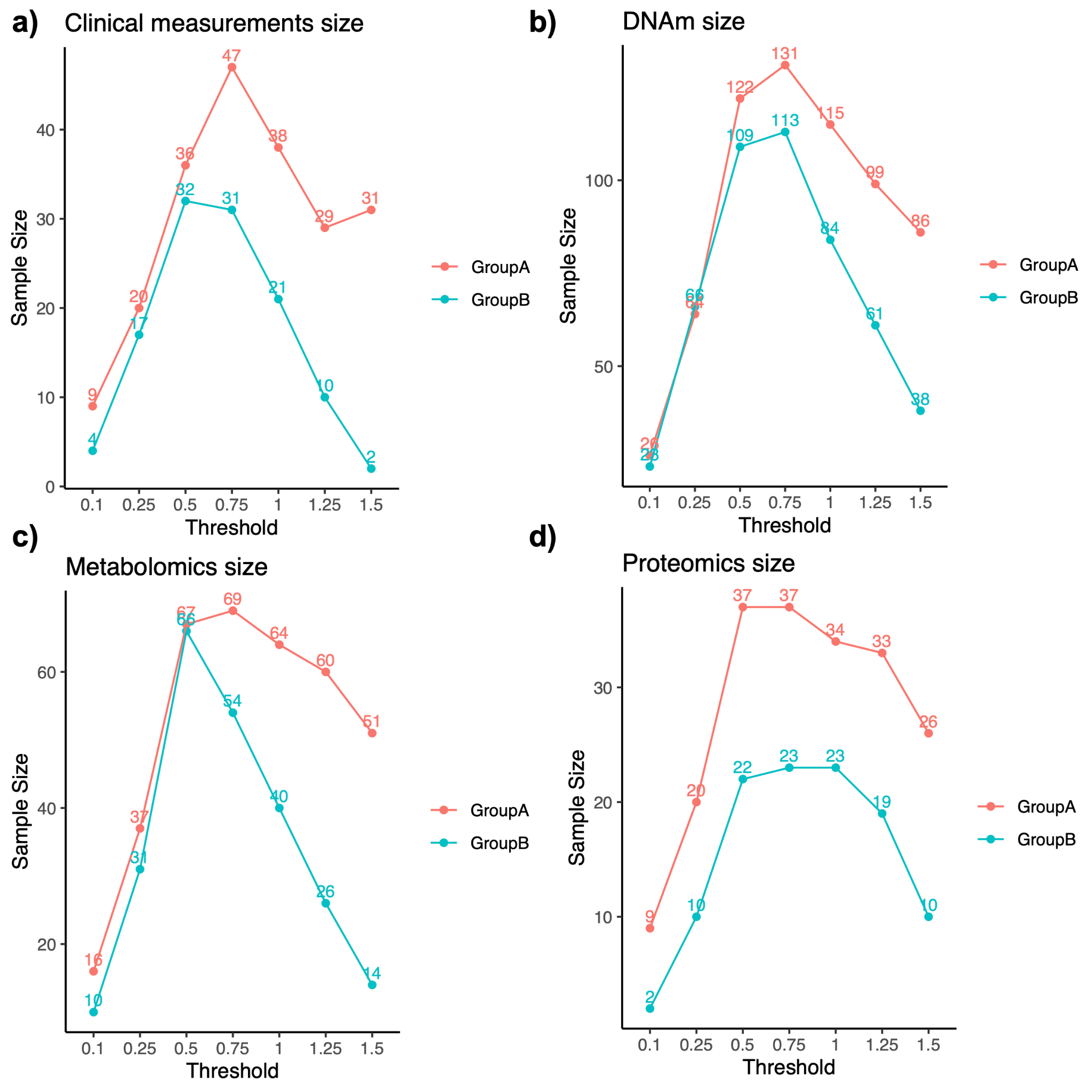

**Figure S1 Number of samples with different threshold of grouping.** **a)** Line plot showing the number of samples (y-axis) in Group A and Group B, when twins are grouped based on whether one twin’s rBMI is above or below ±0.1 SD, ±0.25 SD, ±0.5 SD, ±0.75 SD, ±1 SD, ±1.25 SD, ±1.5 SD of mean, while the co-twin’s rBMI falls within the same threshold. **b)** the same as a), but for DNAm data. **c)** the same as a), but for metabolomics data. **d)** the same as a), but for proteomics data. Group A: BMI-discordant (delta BMI>3) same-sex twin pairs with the higher-BMI twin’s BMI above genetic prediction and the lower-BMI twin’s BMI within genetic prediction; Group B: BMI-discordant (delta BMI>3) same-sex twin pairs with the higher-BMI twin’s BMI within genetic prediction and the lower-BMI twin’s BMI below genetic prediction.

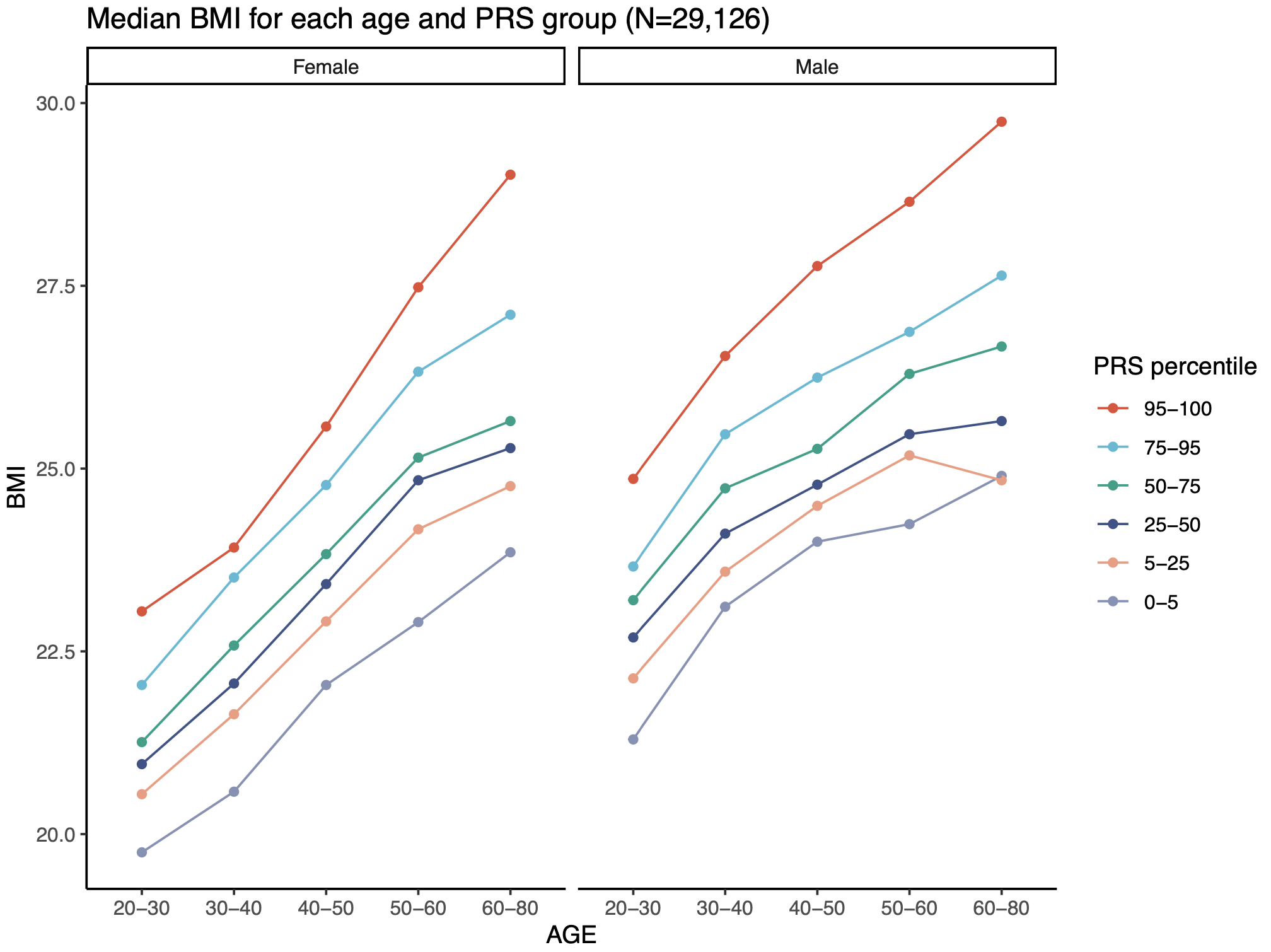

**Figure S2 Interaction of BMI with PRS, Age, and Sex.** Line chart showing the median BMI of individuals across different age groups (x-axis) and PRS percentiles (color), using longitudinal BMI measurements from FTC. A total of 29,126 measurements from 8,785 individuals from the old cohort, 1,424 individuals from FT12 cohort, and 1,219 individuals from FT16 cohort were used.

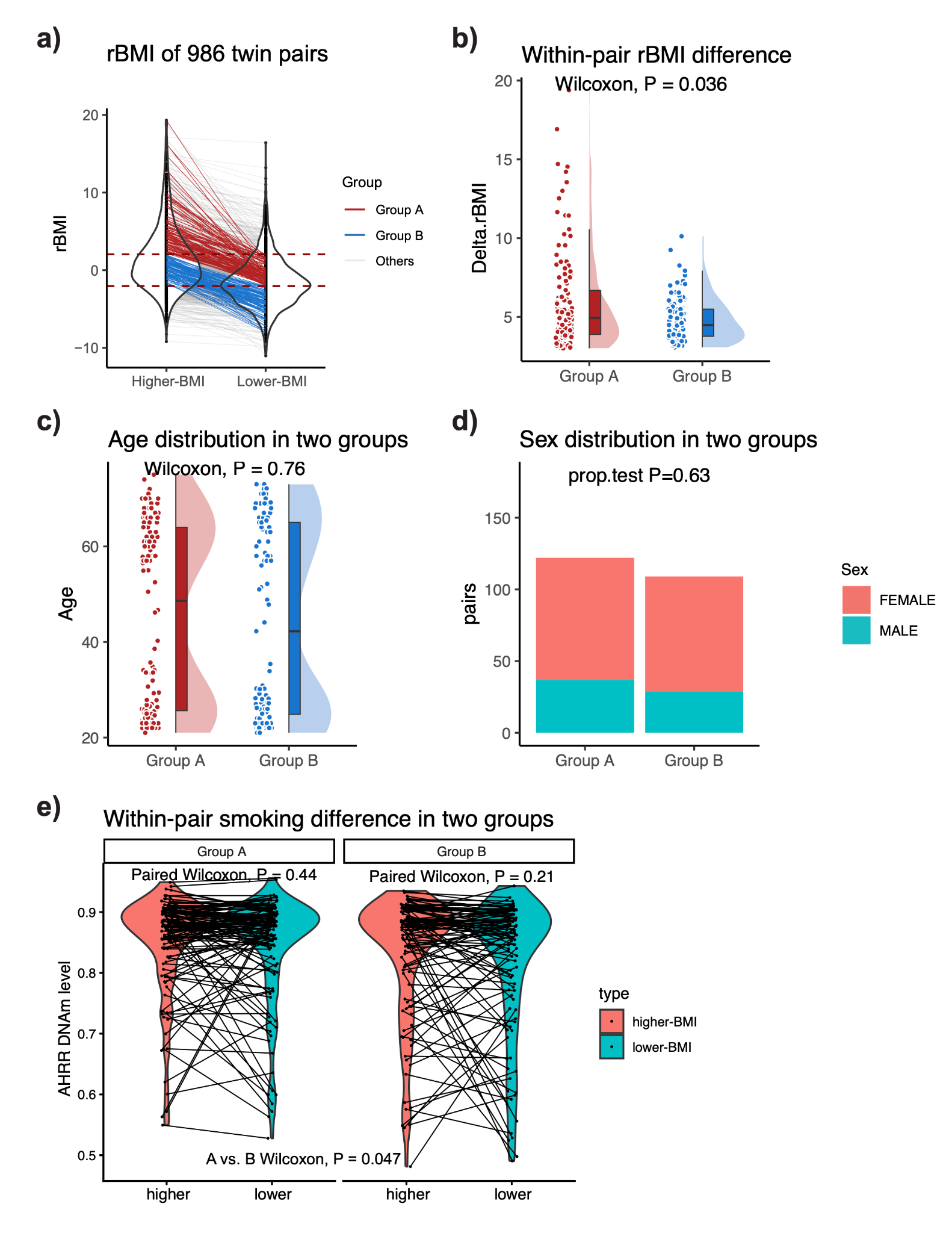

**Figure S3 Within-pair rBMI difference, age, sex and smoking distribution in two groups.** **a)** Violin plot showing the rBMI of the higher-BMI and the lower-BMI twin in each pair, which is linked with red line for group A (above-prediction), blue line for group B (below-prediction), and grey line for the rest. **b)** Violin plot showing the rBMI difference between higher-BMI twin and lower-BMI twin in each twin pair for two groups. P-value is from the Wilcoxon Test. **c)** Violin plot showing the age distribution in each of the two groups. P-value is from the Wilcoxon Test. **d)** Barplot showing the sex distribution in each of the two groups. P-value is from a proportional test. Group A: BMI-discordant (delta BMI>3) same-sex twin pairs with the higher-BMI twin’s BMI above genetic prediction and the lower-BMI twin’s BMI within genetic prediction; Group B: BMI-discordant (delta BMI>3) same-sex twin pairs with the higher-BMI twin’s BMI within genetic prediction and the lower-BMI twin’s BMI below genetic prediction. **e)** Violin plots showing the smoking levels of the higher-BMI and lower-BMI co-twins in group A and B, with each twin pair connected by a line. Smoking was proxied by DNA methylation levels at AHRR (cg05575921). P-values were derived from paired Wilcoxon tests for higher vs. lower-BMI co-twins comparison, and Wilcoxon test for group A vs. B comparison. Group A: BMI-discordant (delta BMI>3) same-sex twin pairs with the higher-BMI twin’s BMI above genetic prediction and the lower-BMI twin’s BMI within genetic prediction; Group B: BMI-discordant (delta BMI>3) same-sex twin pairs with the higher-BMI twin’s BMI within genetic prediction and the lower-BMI twin’s BMI below genetic prediction.

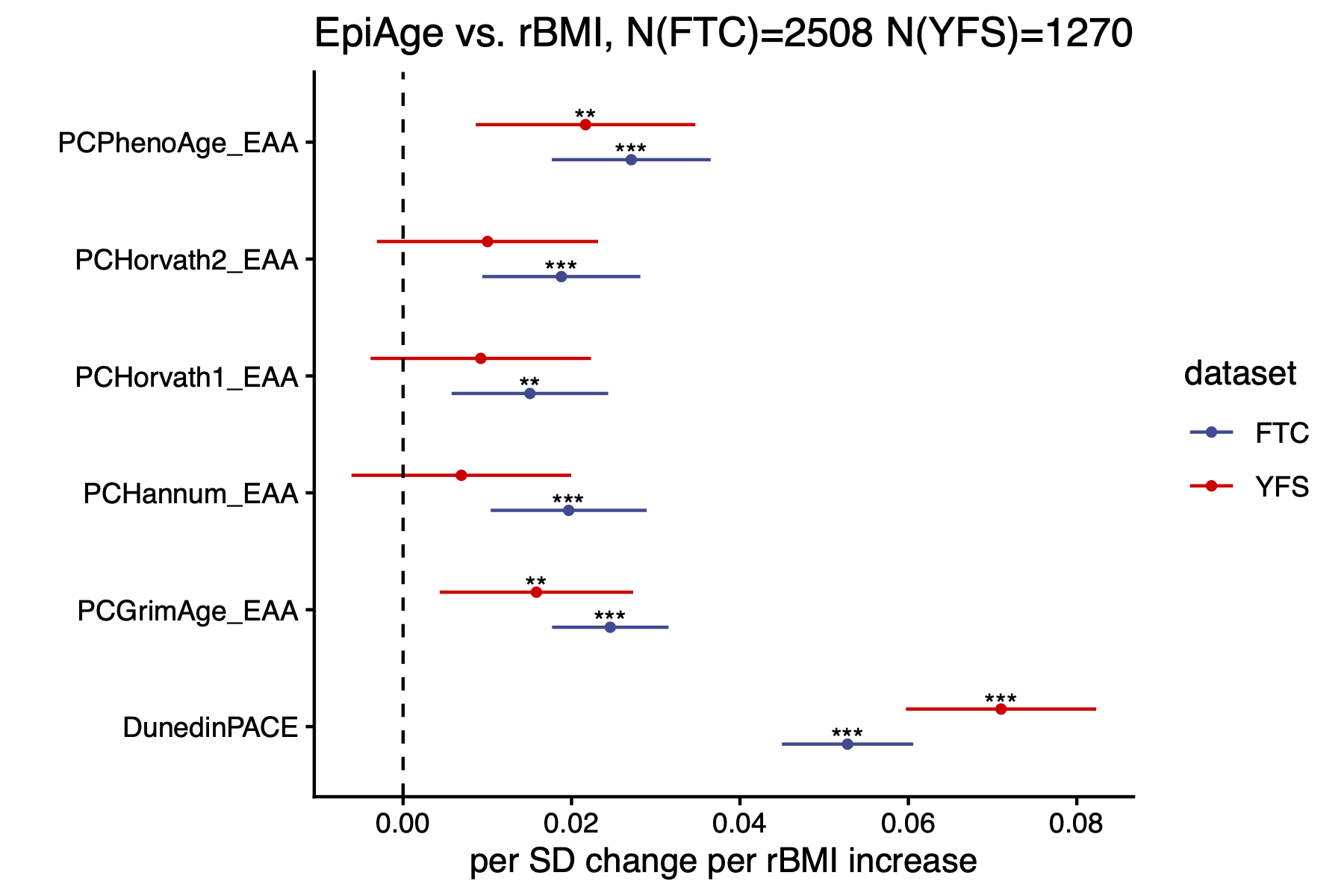

**Figure S4 Correlation between epigenetic age acceleration and rBMI in FTC and YFS participants.** Forest plots display the regression coefficients (dots) and 95% confidence intervals (error bars) for the association between rBMI (x-axis) and epigenetic age acceleration derived from PC clocks and DunedinPACE (y-axis). Epigenetic age acceleration was regressed on rBMI with adjustment for age and sex as fixed effects in both FTC and YFS, and with family ID included as a random effect in FTC. All epigenetic age acceleration measures, including DunedinPACE, were standardized to a mean of 0 and standard deviation of 1. Blue indicates results from 2,508 FTC participants, and red indicates results from 1,270 YFS participants. PCHorvath1 refers to the PC-based Horvath pan-tissue clock, and PCHorvath2 refers to the PC-based Horvath skin and blood clock. Significance levels: *P < 0.05; **P < 0.01; ***P < 0.001; ****P < 0.0001.

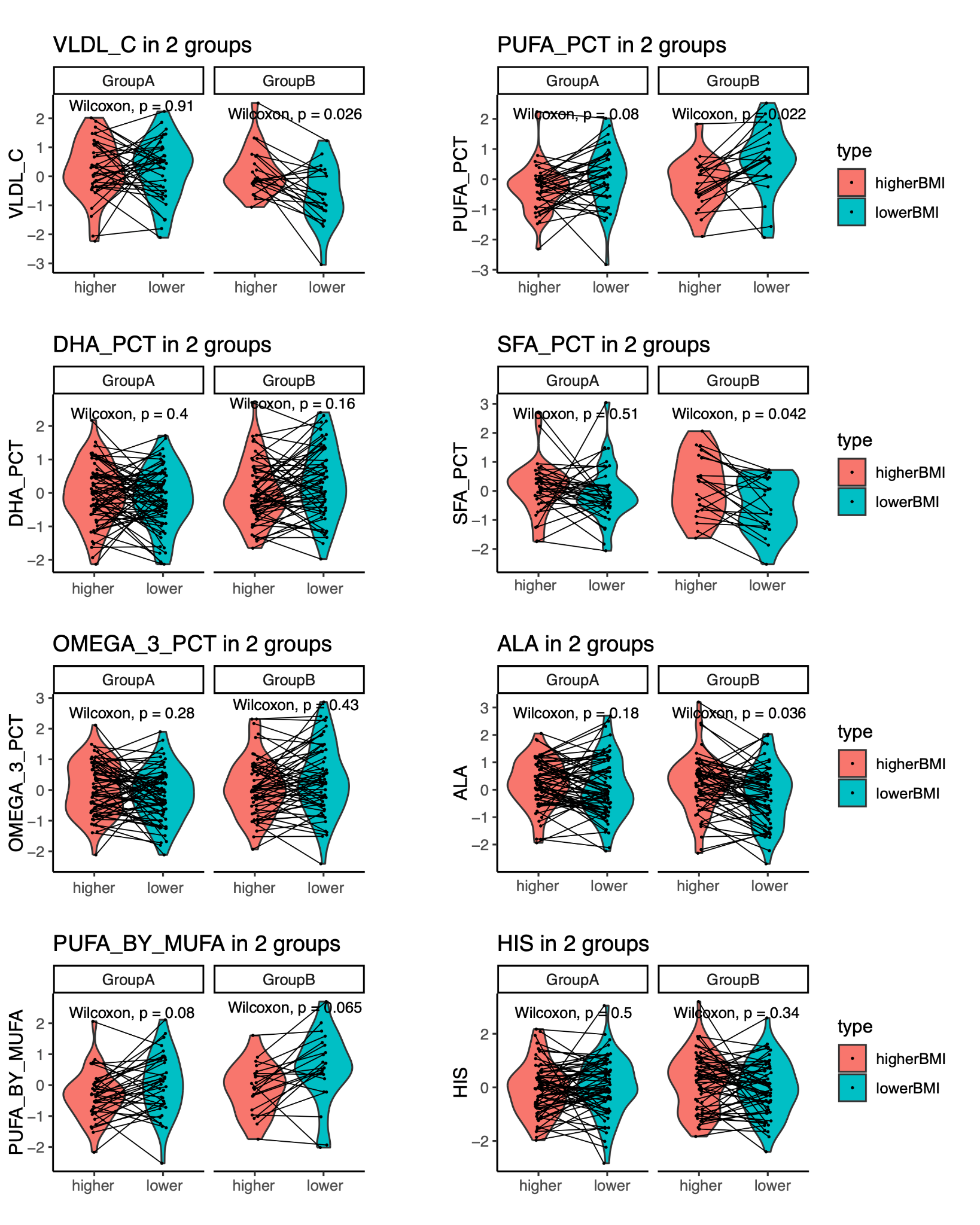

**Figure S5** **Metabolites showing divergent higher- vs. lower-BMI trends between the two groups.** Boxplot showing normalized metabolites levels (y-axis) of higher- and lower-BMI co-twins (x-axis) in the two groups, These metabolites exhibit significant differences between the two groups (A-B) difference with P<0.05). Lines connect co-twins in the same twin pair. P-values were derived from paired Wilcoxon Rank Sum Test. Group A: BMI-discordant (delta BMI>3) same-sex twin pairs with the higher-BMI twin’s BMI above genetic prediction and the lower-BMI twin’s BMI within genetic prediction; Group B: BMI-discordant (delta BMI>3) same-sex twin pairs with the higher-BMI twin’s BMI within genetic prediction and the lower-BMI twin’s BMI below genetic prediction.

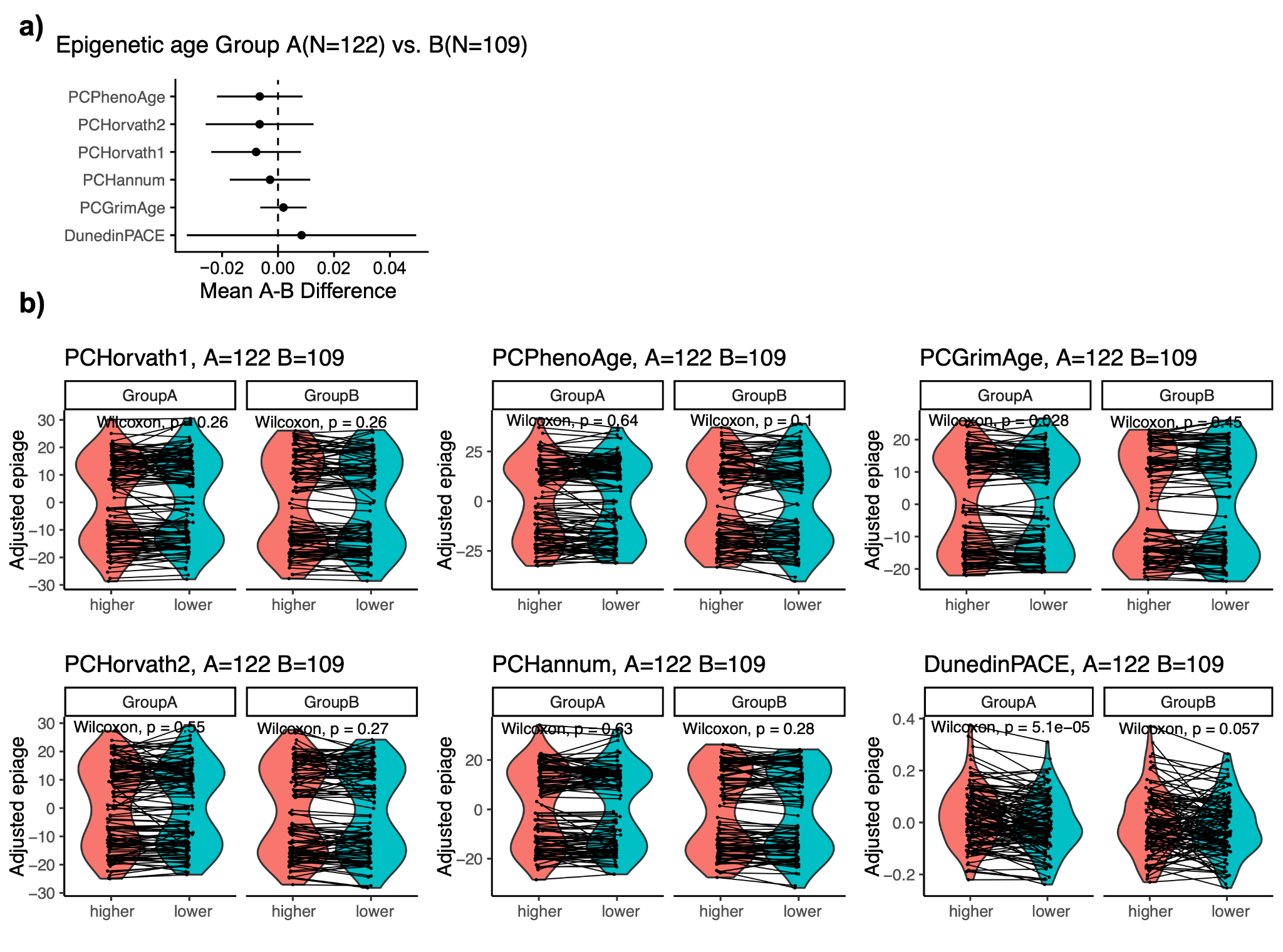

**Figure S6 Epigenetic age of higher- and lower-BMI twins in two groups. a)** Dot plot showing the coefficients (dots) and 95% confidence intervals (error bars) derived from fixed-effect within-pair linear regression using twin pairs from the two groups, where epigenetic age (y-axis) were regressed against rBMI interacting with the group variable and adjusted for smoking. **b)** Boxplot showing epigenetic age (y-axis, adjusted for smoking) of higher- and lower-BMI co-twins (x-axis) in the two groups. Lines connect co-twins in the same twin pair. P-values were derived from paired Wilcoxon Rank Sum Test. Group A: BMI-discordant (delta BMI>3) same-sex twin pairs with the higher-BMI twin’s BMI above genetic prediction and the lower-BMI twin’s BMI within genetic prediction; Group B: BMI-discordant (delta BMI>3) same-sex twin pairs with the higher-BMI twin’s BMI within genetic prediction and the lower-BMI twin’s BMI below genetic prediction.

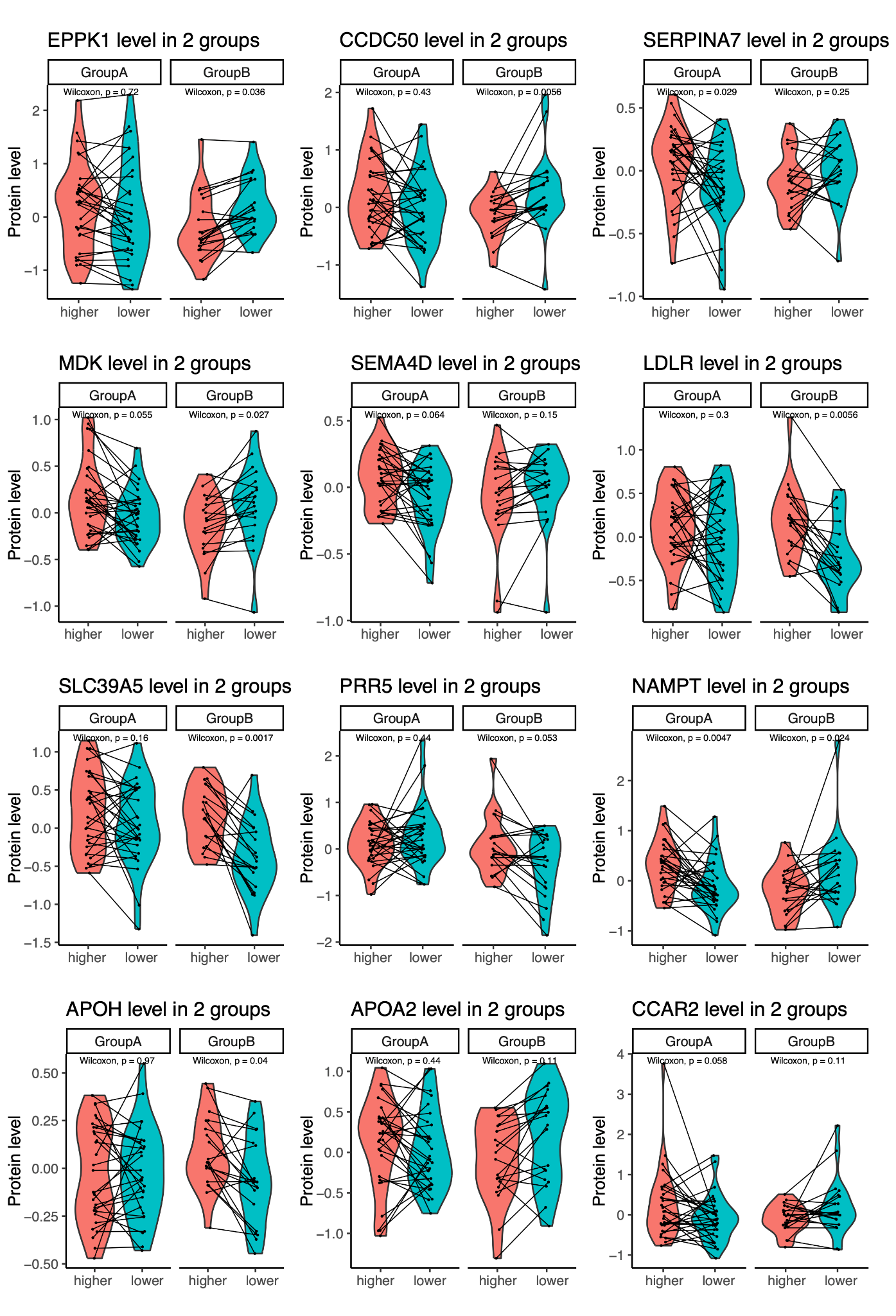

**Figure S7 Top 12 proteins with the largest differences in expression-rBMI correlation between Group A and B.** Boxplot showing normalized protein expression level (y-axis) of higher- and lower-BMI co-twins (x-axis) in each group. Lines connect co-twins in the same twin pair. P-values were derived from paired Wilcoxon Rank Sum Test. Group A: BMI-discordant (delta BMI>3) same-sex twin pairs with the higher-BMI twin’s BMI above genetic prediction and the lower-BMI twin’s BMI within genetic prediction; Group B: BMI-discordant (delta BMI>3) same-sex twin pairs with the higher-BMI twin’s BMI within genetic prediction and the lower-BMI twin’s BMI below genetic prediction.

**Supplementary Methods**

**Preprocessing of FTC genotype data**

This genotyping and quality control pipeline follows the procedures described by ^1^. Genotyping was performed using Illumina Human610-Quad v1.0 B, Human670-QuadCustom v1.0 A, several versions of the Illumina HumanCoreExome arrays (12 v1.0 A, 12 v1.1 A, 24 v1.0 A, 24 v1.1 A, 24 v1.2 A), and the Affymetrix FinnGen Axiom array. Genotypes were called using GenCall for HumanCoreExome arrays, Illuminus for the 610k and 670k arrays, and AxiomGT1 for the Affymetrix array. Quality control (QC) was conducted separately for three batches (batch 1: 610k + 670k, batch 2: HumanCoreExome, batch 3: Affymetrix) following standard procedures. Variants were excluded if their call rate was below 97.5% (batches 1 and 3) or 95% (batch 2), minor allele frequency was below 1%, or Hardy–Weinberg equilibrium P<1×10^-6^. Samples were removed if their call rate was below 98% (batch 1) or 95% (batches 2 and 3), if they failed sex checks, showed excessive heterozygosity (F < -0.03 or F > 0.05 for batches 1 and 2, or ±4 SD from the mean for batch 3), or were identified as population outliers based on multidimensional scaling. After QC, the number of autosomal variants retained was 475,526 (batch 1), 239,894 (batch 2), and 388,673 (batch 3), with 2,617, 5,328, and 8,218 samples, respectively, available for imputation.

**Preprocessing of FTC DNA methylation data**

The DNA methylation data were pre-processed using the *meffil* R package^2^ and details can be found in Sehovic et al.^3^. Low-quality samples were excluded based on sex mismatch, extreme signal deviations, failed control probe metrics, or >20% of probes with detection P-value>0.05 or bead count<3. Functional normalization including the control probe principal components was performed, followed by probe filtering (detection p value > 0.05 or bead number<3 in >20% of samples, sex chromosomes, cross-reactive/polymorphic CpGs^4,5^). BMIQ normalization implemented in the R package *wateRmelon*^6^ was then performed to adjust for type 2 probe bias. The same pipeline was used for 450K and EPIC data. Outliers (beta values deviating >0.2 from nearest neighbors or exceeding 5× the 3rd largest difference) were set to NA, and CpGs with >10% missingness were excluded. Final datasets included 390,304 probes times 1,714 samples 450K data matrix, and 765,385 probes times 1,326 samples EPIC data matrix.

**Preprocessing of FTC metabolomics data**

Metabolite quantification for OLD cohort and FT12 samples was performed using high-throughput ^1^H-NMR (Nightingale Health Ltd, Helsinki, Finland) ^7–9^, as described previously ^10^. Data processing, detailed in ^11^, included 1) exclusion of pregnant women and individuals on cholesterol-lowering medications; 2) imputation of missing values (<10% NA) with sample minimum and excluding metabolites with >10% NA; and 3) Outlier removal (participants with principle components >5 SD from the mean). Metabolites were inverse normal rank transformed (mean = 0, 1 unit = 1 SD). The final merged dataset contains 411 OLD cohort and 657 FT12 samples and 238 metabolites.

**Preprocessing of OLD cohort proteomics data**

415 plasma samples (120 mL each) were analyzed using Olink Explore 3072 comprising cardiometabolic (I/II), inflammation (I/II), neurology (I/II) and oncology (I/II) panels (Olink Proteomics AB, Uppsala, Sweden). Assays failing quality control (e.g., BMP6, EPHX2, PGLYRP1, etc.; full list in ^12^) were excluded. Data were generated as Normalised Protein eXpression (NPX) values. Proteins detected in less than 80% of the samples were excluded. NPX values below limit of detection (LoD) were replaced with the LoD value. Outliers were removed if they met any of the three criteria: (1) PCA scores >5 SD from the mean for the first or the second principle component, (2) median NPX >5 SD from the mean median, or (3) NPX interquantile range >5 SD from the mean IQR. After QC, 2,321 proteins remained; missing values for 17 proteins were imputed using their minimum LoD (average missing value rate for these proteins: 0.5%). The final dataset included 202 same-sex twin pairs.

**Preprocessing of YFS genotype data**

This genotyping and quality control pipeline follows the procedures described by ^13^.

Genotyping was performed at the Wellcome Trust Sanger Institute using a custom-built Illumina Human 670k Custom BeadChip. Initial genotype calling was executed with the Illuminus clustering algorithm. Quality control (QC) was conducted using standard protocols: markers were excluded if they had a call rate <95%, minor allele count (MAC) <1, or significantly deviated from Hardy-Weinberg equilibrium (P<1x10-6). Samples were removed based on high missingness (>5%), gender discrepancies, or high identity-by-descent indicating cryptic relatedness (pi-hat>0.2). To expand the genomic coverage, genotypes were phased and imputed on Michigan Imputation Server using Eagle and SHAPEIT (chromosome X) for phasing and Minimac3 for imputation with the HRC r1-1 reference panel.

**Preprocessing of YFS DNAm data**

Illumina .idat files were processed with the minfi Bioconductor package v1.46.0(Aryee et al., 2014). Probes with SNPs were removed using the dropLociWithSnps function from minfi with default setting. We further removed cross-reactive probes and probes on chromosome X and Y(McCartney et al., 2016; Pidsley et al., 2016). Then, we used the Illumina definition of beta-values and derived P-values of detection for the rest of probes by comparing the total intensity U+M to that of the background distribution (given by negative control probes), as implemented in minfi. Beta-values with P-values of detection greater than 0.05 were set to NA. We retained samples with less than 5% missing values and probes with less than 1% missing values. The missing beta values were then imputed with the impute.knn function (using k = 5) in R. Type-2 probe bias was corrected using BMIQ(Teschendorff et al., 2013). All this resulted in a 770,196 probes times 1,445 samples data matrix.

**Preprocessing of YFS metabolomics data**

Circulating metabolic measures were quantified using a high-throughput nuclear magnetic resonance (NMR) metabolomics platform ^9,14^. Raw data were processed as described previously ^15^. Metabolites were inverse normal rank transformed (mean = 0, 1 unit = 1 SD). The final dataset includes 1715 samples and 158 metabolites.

**Association between PRS for BMI and clinical and metabolic measurements in FTC**

We used linear mixed-effect regression models, with each clinical or metabolic measurement as the dependent variable and PRS for BMI included as the independent variable. Descriptions of clinical and metabolic measurements are provided in the Supplementary Methods and in subsequent sections. Age and sex were included as fixed effect covariates, and twin family ID was included as a random intercept to account for relatedness of twins in a pair. All measurements were standardized to a mean of 0 and a standard deviation of 1 prior to analysis.

### **Comparison of rBMI-associated below- and above-prediction effects at population level**

To determine whether rBMI has differential effects on the outcome $y$ (clinical measurements, metabolomics, proteomics, DNAm, or epigenetic clocks) when it is above versus below zero (i.e., when the observed BMI is higher or lower than the genetically predicted BMI), we implemented a hinge regression model on FTC and YFS, with family ID as random intercept to adjust for twin relatedness in FTC. This approach estimates separate slopes on either side of the knot at rBMI = 0. The model was specified as:

$y=\beta_{0}+\beta_{1}\cdot rBMI+\beta_{2}\cdot hinge+covariates+\left( 1 \right| family)$,

where the hinge term is defined as:

$$hinge=max(0,rBMI)$$

In this parameterization, $\beta_{1}$ represents the slope for rBMI values below zero (below-prediction), whereas $\beta_{1}+\beta_{2}$ corresponds to the slope for rBMI values above zero (above-prediction). The statistical significance of $\beta_{2}$ indicates whether the two slopes differ significantly from each other. P values were adjusted for multiple testing using the Benjamini-Hochberg false discovery rate (FDR) procedure, applied separately within each omics layer in each cohort; unless otherwise specified, nominal P values are reported in the main results. Details of hinge regression analyses for each omics layer are described in the Supplementary Methods.

Clinical measurements: We analyzed the association between 18 obesity-related clinical measurements (inflammation, cardiovascular function, fat metrics, insulin resistance, and liver function markers) and rBMI in 392 individuals from the FTC metabolic sub-cohort ^16^. Sample sizes varied among measurements, as summarized in Table S2. All measurements were standardized to a mean of 0 and a standard deviation of 1 prior to analysis. Age and sex were included as covariates.

Metabolomics: We tested associations between 238 metabolites levels and rBMI in 1,138 FTC individuals. We focused our primary analysis on a subset of 39 clinically validated metabolites from Nightingale Health’s NMR metabolomics platform ^17^. These markers represent key metabolic pathways and are certified for diagnostic use. All measurements were standardized to a mean of 0 and a standard deviation of 1 prior to analysis. Age and sex were included as covariates. The same analytical approach was applied to 1,715 YFS participants for the overlapping metabolites.

DNA methylation: We analyzed associations between DNAm levels at 344,235 CpGs measured using 450K and EPIC arrays and rBMI in 2,508 FTC participants across four subsets (OLD cohort EPIC platform, OLD cohort 450K platform, FT12 and FT16 EPIC platform, and FT12 and FT16 450K platform) and 1,270 YFS participants (EPIC platform). Association analyses were conducted separately within each dataset, followed by meta-analysis using an inverse-variance-weighted approach implemented in METAL ^18^ to account for batch effects between datasets while maximizing sample size. To adjust for cell-type heterogeneity, proportions of 12 blood cell-types were included as covariates, estimated using the EpiDISH algorithm ^19^ with a 12-cell-type reference panel ^20^. Smoking was additionally adjusted for using methylation at cg05575921 as a proxy, a CpG site known to be hypomethylated in heavy smokers ^21^. All models further included age, sex, baseline DNAm level (mean beta value per sample), beadchip processing date, and row position as covariates.

Epigenetic clocks: We computed six epigenetic clocks for 2,508 FTC and 1,270 YFS participants, including the PC clocks ^22^ of Horvath pan-tissue clock, Horvath skin and blood clock, Hannum’s clock, GrimAge, PhenoAge, as well as DunedinPACE ^23^. For the PC clocks, we used epigenetic age acceleration (residuals after regressing each clock on chronological age). For DunedinPACE, the clock value itself was used as the outcome. All the age acceleration or clock scores were standardized to a mean of 0 and a standard deviation of 1 prior to analysis. Sex and smoking were included as covariates, the latter proxied by methylation at cg05575921. Chronological age was additionally included as a covariate in models of DunedinPACE to account for residual age-related variation.

Proteomics: We analyzed associations between 2,321 plasma proteins measured using the Olink platform and rBMI for 381 individuals. Normalised Protein eXpression (NPX) values generated by Olink were used. Age and sex were included as covariates. Proteins exhibiting P<0.05 were carried forward for pathway enrichment analysis. KEGG pathway enrichment was performed using the *enrichKEGG* function from the *clusterProfiler* R package with default settings ^24^. The input list consisted of 342 proteins nominally associated with rBMI, and enrichment was evaluated using a hypergeometric test by comparing this list to the default KEGG background defined in clusterProfiler. P-values were adjusted using the Benjamini-Hochberg false discovery rate (FDR).

**References**

1.	Kujala, U. M. *et al.* Polygenic Risk Scores and Physical Activity. *Med. Sci. Sports Exerc.* **52**, 1518–1524 (2020).

2.	Min, J. L., Hemani, G., Davey Smith, G., Relton, C. & Suderman, M. Meffil: efficient normalization and analysis of very large DNA methylation datasets. *Bioinformatics* **34**, 3983–3989 (2018).

3.	Sehovic, E. *et al.* DNA methylation sites in early adulthood characterised by pubertal timing and development: a twin study. *Clin. Epigenetics* **15**, 181 (2023).

4.	Chen, Y. *et al.* Discovery of cross-reactive probes and polymorphic CpGs in the Illumina Infinium HumanMethylation450 microarray. *Epigenetics* **8**, 203–209 (2013).

5.	Zhou, W., Laird, P. W. & Shen, H. Comprehensive characterization, annotation and innovative use of Infinium DNA methylation BeadChip probes. *Nucleic Acids Res.* gkw967 (2016) doi:10.1093/nar/gkw967.

6.	Pidsley, R. *et al.* A data-driven approach to preprocessing Illumina 450K methylation array data. *BMC Genomics* **14**, 293 (2013).

7.	Bogl, L. H. *et al.* Abdominal obesity and circulating metabolites: A twin study approach. *Metabolism* **65**, 111–121 (2016).

8.	Rose, R. J. *et al.* FinnTwin12 Cohort: An Updated Review. *Twin Res. Hum. Genet.* **22**, 302–311 (2019).

9.	Soininen, P., Kangas, A. J., Würtz, P., Suna, T. & Ala-Korpela, M. Quantitative Serum Nuclear Magnetic Resonance Metabolomics in Cardiovascular Epidemiology and Genetics. *Circ. Cardiovasc. Genet.* **8**, 192–206 (2015).

10.	Whipp, A. M., Heinonen-Guzejev, M., Pietiläinen, K. H., Van Kamp, I. & Kaprio, J. Branched-chain amino acids linked to depression in young adults. *Front. Neurosci.* **16**, 935858 (2022).

11.	Drouard, G. *et al.* Multi‐Omic Associations of Epigenetic Age Acceleration Are Heterogeneously Shaped by Genetic and Environmental Influences. *Aging Cell* e70088 (2025) doi:10.1111/acel.70088.

12.	Drouard, G. *et al.* Twin Study Provides Heritability Estimates for 2321 Plasma Proteins and Assesses Missing SNP Heritability. *J. Proteome Res.* **24**, 2689–2697 (2025).

13.	Juhola, J. *et al.* Childhood Physical, Environmental, and Genetic Predictors of Adult Hypertension: The Cardiovascular Risk in Young Finns Study. *Circulation* **126**, 402–409 (2012).

14.	Soininen, P. *et al.* High-throughput serum NMR metabonomics for cost-effective holistic studies on systemic metabolism. *The Analyst* **134**, 1781 (2009).

15.	Mäkinen, V.-P. *et al.* Longitudinal metabolomics of increasing body-mass index and waist-hip ratio reveals two dynamic patterns of obesity pandemic. *Int. J. Obes.* **47**, 453–462 (2023).

16.	Van Der Kolk, B. W. *et al.* Molecular pathways behind acquired obesity: Adipose tissue and skeletal muscle multiomics in monozygotic twin pairs discordant for BMI. *Cell Rep. Med.* **2**, 100226 (2021).

17.	Julkunen, H. *et al.* Atlas of plasma NMR biomarkers for health and disease in 118,461 individuals from the UK Biobank. *Nat. Commun.* **14**, 604 (2023).

18.	Willer, C. J., Li, Y. & Abecasis, G. R. METAL: fast and efficient meta-analysis of genomewide association scans. *Bioinformatics* **26**, 2190–2191 (2010).

19.	Teschendorff, A. E., Breeze, C. E., Zheng, S. C. & Beck, S. A comparison of reference-based algorithms for correcting cell-type heterogeneity in Epigenome-Wide Association Studies. *BMC Bioinformatics* **18**, 105 (2017).

20.	Luo, Q. *et al.* A meta-analysis of immune-cell fractions at high resolution reveals novel associations with common phenotypes and health outcomes. *Genome Med.* **15**, 59 (2023).

21.	Zeilinger, S. *et al.* Tobacco Smoking Leads to Extensive Genome-Wide Changes in DNA Methylation. *PLoS ONE* **8**, e63812 (2013).

22.	Higgins-Chen, A. T. *et al.* A computational solution for bolstering reliability of epigenetic clocks: implications for clinical trials and longitudinal tracking. *Nat. Aging* **2**, 644–661 (2022).

23.	Belsky, D. W. *et al.* DunedinPACE, a DNA methylation biomarker of the pace of aging. *eLife* **11**, e73420 (2022).

24.	Yu, G., Wang, L.-G., Han, Y. & He, Q.-Y. clusterProfiler: an R Package for Comparing Biological Themes Among Gene Clusters. *OMICS J. Integr. Biol.* **16**, 284–287 (2012).
